# An integrative analysis of clinical and epigenetic biomarkers of mortality

**DOI:** 10.1101/2022.01.24.22269611

**Authors:** Tianxiao Huan, Steve Nguyen, Elena Colicino, Carolina Ochoa-Rosales, W. David Hill, Jennifer A. Brody, Mette Soerensen, Yan Zhang, Antoine Baldassari, Mohamed Ahmed Elhadad, Tanaka Toshiko, Yinan Zheng, Arce Domingo-Relloso, Dong Heon Lee, Jiantao Ma, Chen Yao, Chunyu Liu, Shih-Jen Hwang, Roby Joehanes, Myriam Fornage, Jan Bressler, Joyce BJ van Meurs, Birgit Debrabant, Jonas Mengel-From, Jacob Hjelmborg, Kaare Christensen, Pantel Vokonas, Joel Schwartz, Sina A. Gahrib, Nona Sotoodehnia, Colleen M. Sitlani, Sonja Kunze, Christian Gieger, Annette Peters, Melanie Waldenberger, Ian J. Deary, Luigi Ferrucci, Yishu Qu, Philip Greenland, Donald M Lloyd-Jones, Lifang Hou, Stefania Bandinelli, Trudy Voortman, Brenner Hermann, Andrea Baccarelli, Eric Whitsel, James S. Pankow, Daniel Levy

## Abstract

DNA methylation (DNAm) has been reported to be associated with many diseases and mortality. We hypothesized that the integration of DNAm with clinical risk factors would improve mortality prediction.We performed an epigenome-wide association study of whole blood DNAm in relation to mortality in 15 cohorts (n=15,013). During a mean follow-up of 10 years, there were 4314 deaths from all-causes including 1235 cardiovascular disease (CVD) deaths and 868 cancer deaths. Ancestry-stratified meta-analysis of all-cause mortality identified 163 CpGs in European ancestry (EA) and 17 in African ancestry (AA) participants at *P*<1x10^-7^, of which 41 (EA) and 16 (AA) were also associated with CVD death, and 15 (EA) and 9 (AA) with cancer death. We built DNAm-based prediction models for all-cause mortality that predicted mortality risk independent of clinical risk factors. The mortality prediction model trained by integrating DNAm with clinical risk factors showed a substantial improvement in prediction of cancer death with 11% and 5% increase in the C-index in internal and external replications, compared with the model trained by clinical risk factors alone. Mendelian randomization identified 15 CpGs in relation to longevity, CVD, or cancer risk. For example, cg06885782 (in *KCNQ4*) was positively associated with risk for prostate cancer (Beta=1.2, *P_MR_*=4.1x10^-4^), and negatively associated with longevity (Beta=-1.9, *P_MR_*=0.02). Pathway analysis revealed that genes associated with mortality-related CpGs are enriched for immune and cancer related pathways. We identified replicable DNAm signatures of mortality and demonstrated the potential utility of CpGs as informative biomarkers for prediction of mortality risk.

## Introduction

Despite substantial evidence of heritability of human longevity (*h*^2^ = 10-30%), genome-wide association studies (GWAS) have reported few loci associated with human longevity (Deelen et al., 2019; Pilling et al., 2017; Timmers et al., 2019; van den Berg, Beekman, Smith, Janssens, & Slagboom, 2017). DNA methylation (DNAm), the covalent binding of a methyl group to the 5’ carbon of cytosine-phosphate-guanine (CpG) dinucleotide sequences, reflects a wide range of environmental exposures and genetic influences at the molecular level and altered DNAm has been shown to regulate gene expression (Jones & Takai, 2001). Recent studies have reported DNAm patterns associated with age in humans (Hannum et al., 2013; Horvath, 2013; Levine et al., 2018; Lu et al., 2019). Estimates of biological age based on DNAm, referred to as "epigenetic age" or "DNAm age" have been validated in numerous studies, although the functions of these age-associated CpGs are largely unknown (Horvath et al., 2015; Lu et al., 2019; Marioni, Shah, McRae, Chen, et al., 2015; Marioni, Shah, McRae, Ritchie, et al., 2015). DNAm age also has been shown to be predictive of many age-related diseases and of all-cause mortality (Chen et al., 2016; Dugué et al., 2018; Levine et al., 2018; Lu et al., 2019; Marioni, Shah, McRae, Chen, et al., 2015).

Despite the association of DNAm age with a variety of age-associated outcomes, age-related CpGs are different from those that are most strongly associated with mortality. Relatively few DNAm studies have focused on mortality as the primary outcome (Colicino et al., 2020; Svane et al., 2018; Zhang et al., 2017). Moreover, due to sample size limitations, most DNAm mortality studies have not typically investigated cause-specific mortality such as death due to cardiovascular disease (CVD) and cancer. Additionally, little is known about the prediction performance of DNAm-based mortality models and whether or not such approaches improve mortality prediction above and beyond established clinical risk factors.

We hypothesized that inter-individual variation in DNAm is associated with all-cause mortality risk and with cause-specific mortality, and that we could build models incorporating CpGs that would improve mortality prediction beyond established clinical risk factors. In this study, we report the results of a meta-analysis of epigenome-wide association studies (EWAS) of all-cause mortality and cause-specific mortality including death from CVD and cancer in up to 15,013 individuals from 15 prospective cohort studies in which DNAm was measured in whole blood. We built all-cause mortality risk prediction models using penalized regression and machine learning methods and integrated DNAm and established mortality clinical risk factors and validated the models’ performance. Additionally, using Mendelian randomization, we identified putatively causal CpGs for mortality. Last, we investigated the downstream gene expression and pathway changes of the mortality-related CpGs by testing associations between DNAm and gene expression. **Fig. S1** summarizes the multi-step study design.

## Results

### Study population

**Table 1** presents the major clinical characteristics of the 15,013 study participants including 11,684 European ancestry (EA, mean age 65, 55% women) and 3329 African ancestry (AA, mean age 59, 70% women) participants from 15 cohorts (**Table S1** summarizes additional clinical characteristics). Most studies had fewer than 15 years of mean follow-up (mean values ranged from 6.4 to 13.7 years), except ARIC (mean follow-up of 20.0 years in ARIC EA and 18.6 in ARIC AA participants, respectively). During follow-up of EA participants, 2907 died of any cause, 688 of CVD, and 546 of cancer; among AA participants, 1407 died of any cause, 547 of CVD, and 322 of cancer.

**Table 1:** Clinical characteristics the 15,013 study participants.

| Cohort | Total N | No. of all-cause death | No. of CVD death | No. of cancer death | Time to death / last follow-up years, mean (SD) | Age , mean (SD) | Sex (F, %) | BMI, mean (SD) | Prevalent Diseases |  |  |  |  |  |
| --- | --- | --- | --- | --- | --- | --- | --- | --- | --- | --- | --- | --- | --- | --- |
|  |  |  |  |  |  |  |  |  | Type 2 Diabetes (n) | Coronary Heart Disease (n) | Heart Failure (n) | Stroke (n) | Hypertens ion (n) | Cancer (n) |
| -- European Ancestry |  |  |  |  |  |  |  |  |  |  |  |  |  |  |
| ARIC | 969 | 331 | 95 | 94 | 20.0 (5.2) | 59.8 (5.5) | 59 | 26.2 (4.5) | 86 | 44 | 29 | 16 | 233 | 102 |
| CHS | 419 | 373 | 132 |  | 12.7 (6.1) | 75.0 (4.9) | 60 | 26.8 (4.9) | 72 | 16 | 11 | 5 | 224 | 78 |
| DTR | 870 | 298 | 74 | 40 | 9.3 (3.4) | 69.4 (7.9) | 52 | 25.9 (3.9) | 46* |  |  | 37 | 269 | 129 |
| ESTHER | 1000 | 265 | 94 | 90 | 13.7 (3.5) | 62.1 (6.5) | 50 | 27.8 (4.3) | 154 | 144 | 110 | 28 | 572 | 77 |
| FHS | 2427 | 403 | 91 | 155 | 9.1 (2.2) | 66.3 (9.0) | 55 | 28.3 (5.3) | 279 | 226 | 53 | 116 | 107 | 389 |
| InCHIANTI | 488 | 104 |  |  | 10.0 (1.6) | 62.4 (15.8) | 52 | 27.0 (3.9) | 42 | 31 | 9 | 10 | 232 |  |
| KORA F4 | 1727 | 89 | 31 | 35 | 6.4 (0.9) | 61.0 (8.9) | 51 | 28.1 (4.8) | 158 | 105 | 41 | 47 | 789 | 154 |
| LBC 1921 | 418 | 366 |  |  | 9.8 (4.7) | 79.1 (0.6) | 60 | 28.2 (4.0) | 19 | 70 |  | 33 | 170 |  |
| LBC 1936 | 900 | 192 |  |  | 10.2 (2.4) | 69.6 (0.8) | 50 | 27.7(4.4) | 72 | 221 |  | 46 | 364 |  |
| NAS | 640 | 221 | 123 | 72 | 10.5 (3.3) | 72.8 (6.8) | 0 | 28.1 (4.0) | 117 | 181 |  | 42 | 447 | 316 |
| RS | 731 | 73 |  |  | 6.8 (1.5) | 59.9 (8.2) | 54 | 27.4 (4.5) | 74 | 45 |  | 30 | 385 | 76 |
| WHI | 1095 | 192 | 48 | 60 | 11.5 (3.5) | 62 (6.9) | 100 | 28.8 (5.9) | 60 | 20 | 5 | 11 | 469 | 14 |
| -- African Ancestry |  |  |  |  |  |  |  |  |  |  |  |  |  |  |
| ARIC | 2446 | 1069 | 424 | 322 | 18.6 (6.6) | 56.5 (5.8) | 64 | 30.1 (6.2) | 643 | 120 | 163 | 75 | 1373 | 87 |
| CHS | 325 | 264 | 96 |  | 12.9 (6.6) | 73.1 (5.5) | 62 | 28.6 (5.2) | 68 | 2 | 0 | 2 | 235 | 36 |
| WHI | 558 | 74 | 27 |  | 10.6 (3.7) | 61 (6.8) | 100 | 31.5 (6.1) | 76 | 18 | 11 | 12 | 369 | 2 |

\*The diabetes cases in DTR included both type-I and type-II diabetes.

### Ancestry-stratified epigenome-wide meta-analysis of all-cause mortality

At Bonferroni-corrected *P*<1x10^-7^ (∼0.05/400,000), we identified 163 CpGs whose differential methylation in whole blood was associated with all-cause mortality in EA participants, and 17 CpGs in AA participants, after adjustment of age, sex, lifestyle factors, clinical risk factors, white blood cell types, and technical covariates. **Tables S2-S3** present the results for all CpGs at *P*<1x10^-5^. Overall genomic inflation in meta-analysis (λ) was estimated at 1.15 or less, indicating low inflation and low risk of false-positive findings. Even though cohort-specific analysis showed slightly higher genomic inflation in some cohorts (λ>1.5 in two cohorts, **Table S4**), forest plots show that the results were not driven by results from one or several cohorts (**Fig. S2**). Sensitivity analysis results including meta-analysis after correcting for λ in each cohort, meta-analysis after excluding results from two cohorts with λ >1.5 and meta-analysis after excluding RS cohort are included in **Table S5-S6**. Results of the sensitivity analysis remained consistent with the main results in terms of direction and effect estimates with Pearson’s correlation *r*= 0.99 (in EA, corrected for λ in each cohorts), *r*= 1.00 (in EA, after removing two cohorts with λ >1.5), *r*= 1.00 (in EA, after removing RS) and *r*= 1.00 (in AA, corrected for λ in each cohorts).

Among the 177 all-cause mortality-related CpGs (union set of EA and AA results at *P*<1x10^-7^), the vast majority of significant CpGs (151, 85%) were inversely associated with mortality, with hazards ratios (HRs) <1 (range 0.72 to 0.89 per standard deviation [SD]). Methylation at the remaining 26 (15%) CpGs was positively associated with mortality, with HRs >1 (range 1.13 to 1.32). The 177 CpGs are annotated to 121 genes and 43 intergenic regions.

### Transethnic replication and sensitivity analysis

Of the 163 all-cause mortality related CpGs in EA participants, 18 (11%) had *P*< 0.0003 (0.05/163) in AA participants; of the 17 CpGs in AA participants, 12 (71%) had *P*< 0.004 (0.05/17) in EA participants. **Table 2** displays the transethnic replicated CpGs including 27 unique CpGs. The top 3 transethnic replicated CpGs in EA participants remained the top 3 in AA participants, including cg16743273 for *MOBKL2A*, cg18181703 for *SOCS3*, and cg21393163 at an intergenic region (Chr.1: 12217629).

**Table 2:**
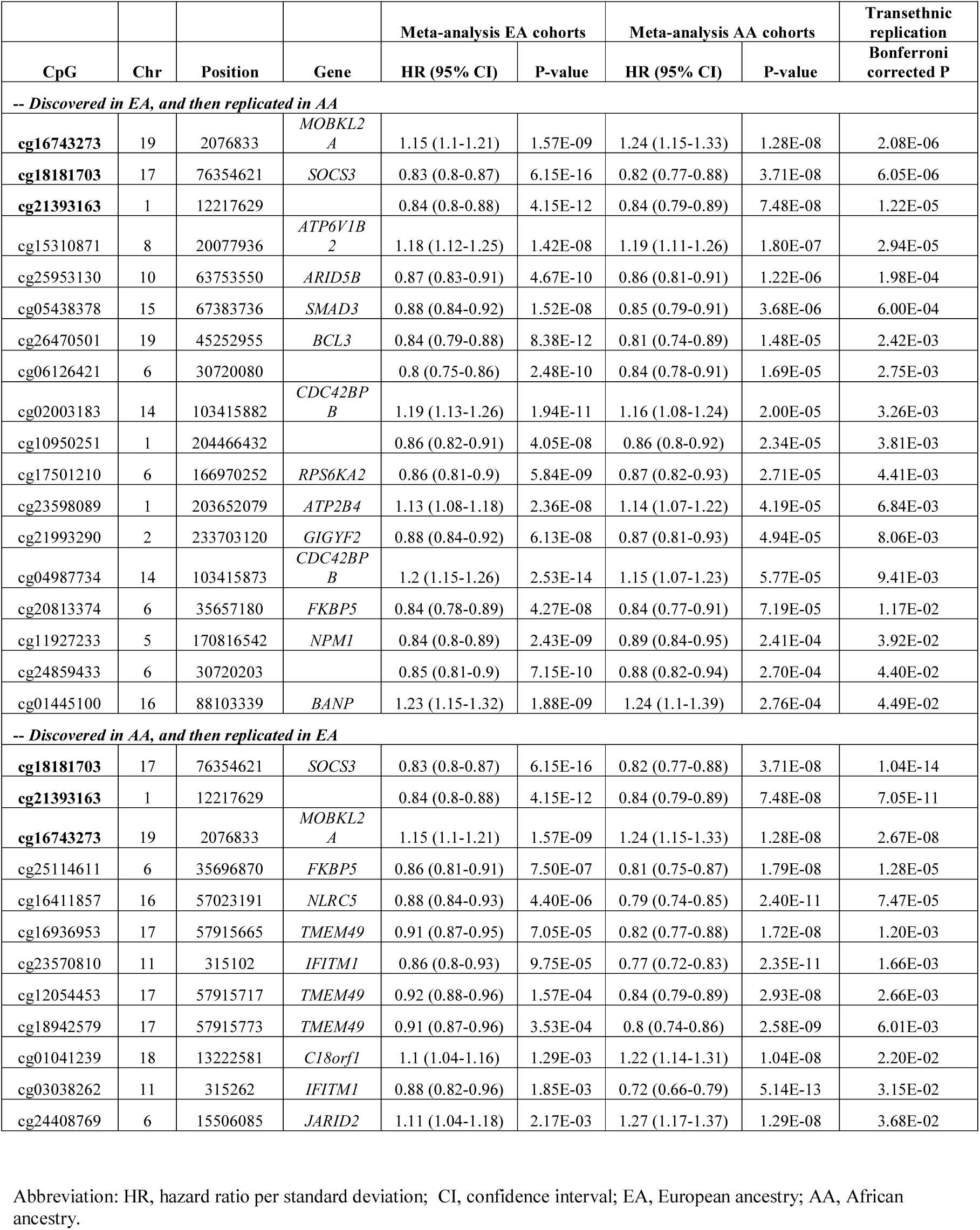
Trans-ethnic replicated all-cause mortality related CpGs.

Because ARIC had longer follow-up than the other cohorts, in sensitivity analysis, we truncated ARIC follow up at 15 years. The HRs for the significant CpGs (at *P*<1x10^-5^) remained consistent with the main results in terms of direction and effect estimates with Pearson’s correlation *r*= 1.00 and *r*= 0.99 in EA and AA participants, respectively (**Table S2-S3** and **Fig. S3**).

### Associations of DNAm with CVD death and cancer death

In comparison with results for all-cause mortality, fewer CpGs were associated with CVD death (at *P*<1x10^-7^, n=4 in EA, and n=15 in AA) and cancer death (n=0 in EA, and n=1 in AA) **Tables S7-S8** report the corresponding results at *P*<1x10^-5^. Among the 163 all-cause mortality-related CpGs identified in EA participants at *P*<1x10^-7^, 41 CpGs were associated with CVD death, 16 with cancer death, and 5 with both (at *P*<0.05/163, **Table S2**). Among the 17 CpGs identified in AA participants at *P*<1x10^-7^, 15 were associated with CVD death, 9 with cancer death, and 8 with both (at *P*<0.05/17, **Table S3**). **Fig. 1** shows the effect sizes and direction of effect for the top CpGs associated with all-cause mortality, and their consistency with the results of analyses of CVD death and cancer death. We found that if a CpG was positively correlated with all-cause mortality, then it also was positively correlated with CVD death and cancer death, and vice versa.

**Figure 1:**
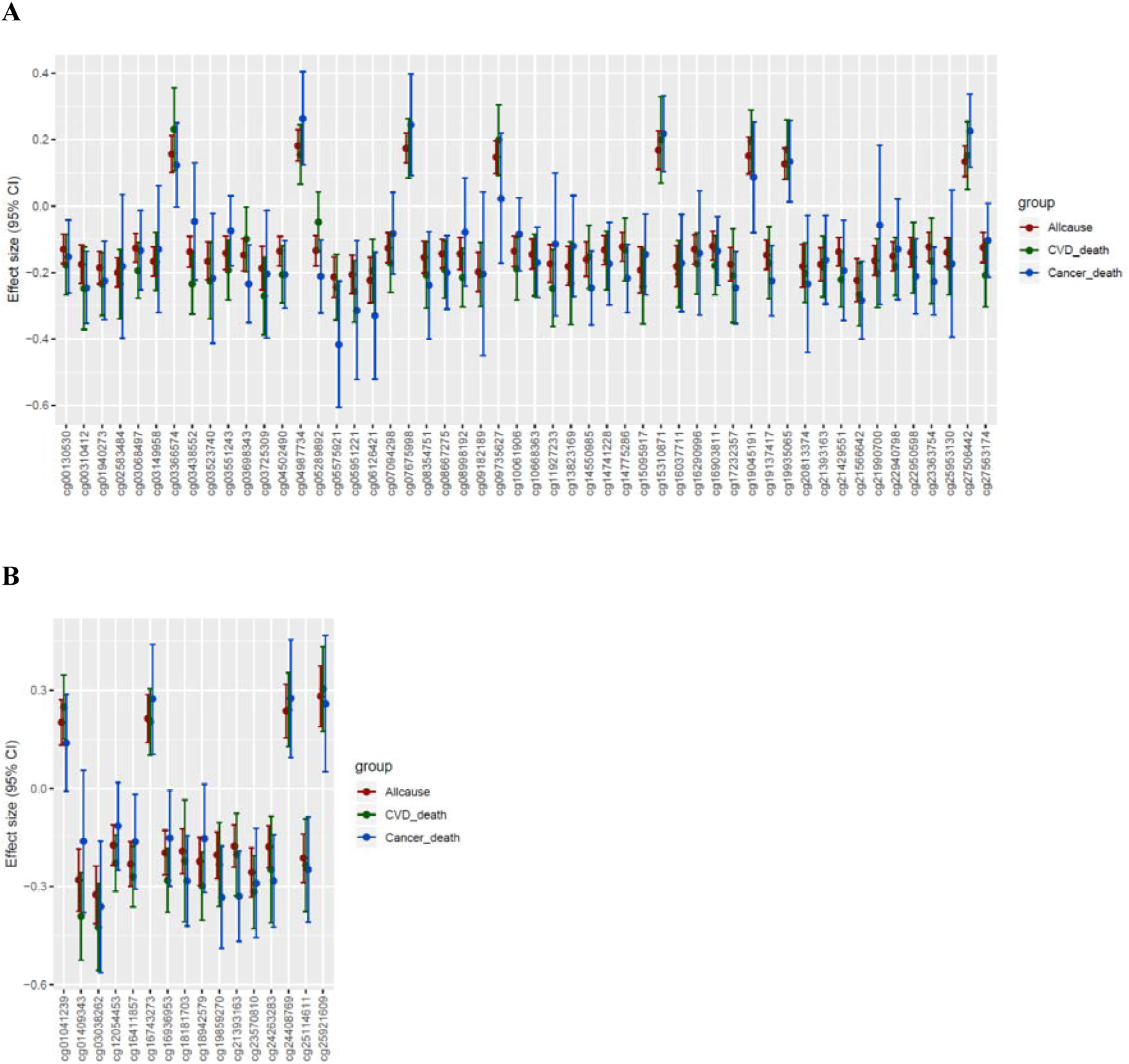
The effect sizes (log hazards ratios) and 95% confidence intervals of CpGs related to mortality identified by meta-analysis, comparing the results for all-cause mortality, CVD death, and cancer death. A) Results of meta-analysis of European ancestry (EA); B) Results of meta-analysis of African ancestry (AA). These figures showed the CpGs associated with all-cause mortality identified by the meta-analysis, which were also associated with either CVD death or cancer death passing Bonferroni corrected threshold. Figure 1A shows 51 CpGs in EA, including 41 CpGs associated with CVD death, 16 with cancer death, and 5 with both. Figure 1B shows 16 CpGs in AA, including 15 CpGs associated with CVD death, 8 with cancer death, and 7 with both.

**Figure 2:**
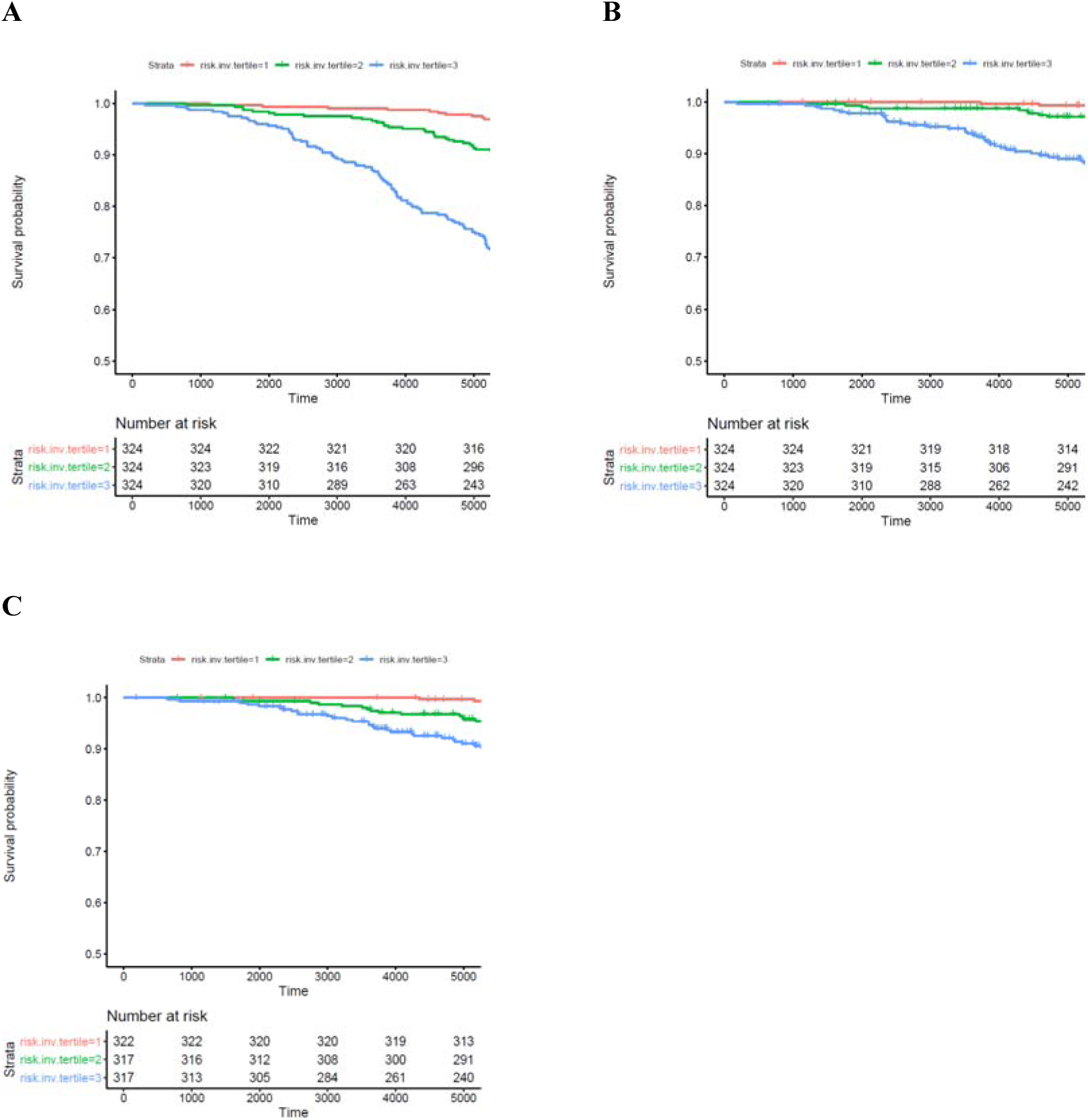
Kaplan–Meier estimates of mortality risk scores with respect to mortality outcomes in ARIC study. A) survival curves with respect to all-cause mortality; B) survival curves with respect to CVD death; C) survival curves with respect to cancer death. The results were obtained from ARIC European ancestry participants with follow up truncated at 15 years. For cancer death, we excluded samples who had any type of cancer before blood drawn for DNA methylation measurements. The mortality risk scores for A) and B) were computed by the model (**Table S10**), and for C) was computed by the model (**Table S11**)

### Mortality prediction model

To investigate if DNAm can be used to predict mortality risk, we constructed prediction models for all-cause mortality, and evaluated their prediction of all-cause mortality, CVD death, and cancer death. To ensure unbiased validation, we split the EA cohorts into separate discovery and replication sets (**Fig. S1** shows the analysis flowchart). The discovery cohorts consisted of 8288 participants (including 2173 deaths from all-causes) from 10 cohorts, excluding FHS (n=2427) and ARIC (n=969), which were used as replication cohorts. The meta-analysis of the discovery set identified 74 CpGs at *P*<1x10^-7^, 158 CpGs at *P*<1x10^-6^, 357 CpGs at *P*<1x10^-5^, 931 CpGs at *P*<1x10^-4^, 2717 CpGs at *P*<1x10^-3^, and 28,323 CpGs at *P*<0.05. We evaluated three types of input features: a) clinical risk factors only (i.e., clinical risk factor models); b) CpGs identified in the meta-analysis of the discovery set (i.e., CpG models); and c) the input features including both CpGs and clinical risk factors (i.e., integrative models). We also compared four prediction methods including Elastic net - Cox proportional hazards (Elastic-coxph) (Friedman, Hastie, & Tibshirani, 2010), Random survival forest (RSF) (Ishwaran, Kogalur, Blackstone, & Lauer, 2008), Cox-nnet (Ching, Zhu, & Garmire, 2018), and DeepSurv (Katzman et al., 2018) (see Methods for details). In general, the four prediction methods did not show major differences in predicting mortality outcomes as assessed by multiple evaluation metrics (**Table S9** lists the evaluation metrics across all four methods). To simplify the presentation of results, we focused on the Elastic-coxph method.

#### Clinical risk factors strongly predict all-cause mortality and CVD death

The C-index of the clinical risk factor models (age, sex, and 12 clinical risk factors) was 0.80 for all-cause mortality, 0.81 for CVD death, and 0.77 for cancer death in FHS (reflecting the average values of 10-fold cross-validation). Among the 12 clinical risk factors, prevalent cancer status was the major contributor to predicting cancer death. After excluding individuals with prevalent cancer at the time of blood draw for DNAm measurements (i.e., the start of follow up), the C-index of the clinical risk factor model was 0.57 for cancer death. Finally, two clinical risk models were built using the optimum parameters selecting by cross-validation (see Methods). The first one was trained using all FHS participants and included 10 risk factors selected by the Elastic-coxph method (to predict all-cause mortality and CVD death, **Table S10**), and the second was trained using FHS participants excluding those with prevalent cancer cases and including 10 risk factors (to predict cancer death, **Table S11**). The corresponding C-index of the clinical risk factor model was 0.75 for all-cause mortality (HR=2.64 per SD in the risk score, 95% CI [2.21, 3.15], *P*=4.4x10^-27^), 0.81 for CVD death (HR=3.51, 95% CI [2.58, 4.79], *P*=2.1 x10^-15^), and 0.71 for cancer death (excluding prevalent cancer samples, HR=2.35, 95% CI [1.74, 3.18], *P*=2.3 x10^-8^) in ARIC EA participants with follow up truncated at 15 years (**Table 3**).

**Table 3:** Performance robustness comparison of mortality predictors in FHS and ARIC cohorts.

|  | FHS* |  |  | ARIC<sup>†</sup> |  |  |
| --- | --- | --- | --- | --- | --- | --- |
| Model | HR | C-index | IBS | HR ( 95% CI) | C-index | IBS |
| *-- All-cause mortality* |  |  |  |  |  |  |
| **Clinical Risk Factor Model** | 3.37 | 0.80 | 0.07 | 2.64 (2.21-3.15) | 0.75 | 0.04 |
| **CpG Model** | 2.91 | 0.77 | 0.07 | 2.24 (1.89-2.66) | 0.72 | 0.04 |
| **Integrative Model** | 3.50 | 0.80 | 0.06 | 2.95 (2.45-3.55) | 0.77 | 0.04 |
| *-- CVD Death* |  |  |  |  |  |  |
| **Clinical Risk Factor Model** | 3.74 | 0.81 | 0.02 | 3.51 (2.57-4.79) | 0.81 | 0.02 |
| **CpG Model** | 3.85 | 0.82 | 0.02 | 2.62 (1.56-3.91) | 0.77 | 0.02 |
| **Integrative Model** | 3.90 | 0.83 | 0.02 | 3.65 (2.63-5.05) | 0.80 | 0.02 |
| *-- Cancer Death (excluding prevalent cancer cases)* |  |  |  |  |  |  |
| **Clinical Risk Factor Model** | 1.25 | 0.57 | 0.01 | 2.35 (1.74-3.18) | 0.71 | 0.02 |
| **CpG Model** | 1.71 | 0.65 | 0.01 | 2.22 (1.64-2.89) | 0.73 | 0.02 |
| **Integrative Model** | 1.78 | 0.68 | 0.01 | 2.58 (1.90-3.50) | 0.76 | 0.02 |

Abbreviation: HR, hazard ratio per standard deviation; IBS: Integrated brier score;
\* HR, C-index and IBS values in FHS reflect the average values of 10 times cross-validation.
<sup>†</sup> The results were obtained from ARIC European ancestry participants with follow up truncated at 15 years.
The clinical risk factor models were trained by using clinical risk factors as the sole input features. The CpG Models were trained by using CpGs selecting in the discovery meta-analysis. The integrative model was trained by using both clinical risk factors and CpGs selecting in the discovery meta-analysis.

#### DNAm predicts mortality independently of age and clinical risk factors

The models using all-cause mortality-related CpGs identified in the discovery cohorts as the sole input feature (the CpG model) were predictive of all-cause mortality, CVD death, and cancer death in the replication set. As shown in **Fig. S4**, when more discovery CpGs were added to the model, the prediction performance metrics did not always improve. In FHS, the models with discovery CpGs at *P*<1x10^-3^ showed the best predictive performance for all-cause mortality (C-index =0.77) and CVD death (C-index =0.82), but the model with discovery CpGs at *P*< 1x10^-5^ showed the best predictive performance for cancer death (excluding prevalent cancer cases, [C-index =0.65]). The final CpG models that were trained using all FHS participants are provided in **Table S12** including 76 CpGs to predict all-cause mortality and CVD death, and in **Table S13** including 56 CpGs to predict cancer death (excluding prevalent cancer cases). The C-index of the CpG models with the best predictive performance in ARIC were 0.72 for all-cause mortality (HR=2.21, 95% CI [1.86, 2.62], *P*=2.0x10^-20^), 0.77 for CVD death (HR=2.62, 95% CI [1.96, 3.51], *P*=9.9x10^-11^) and 0.73 for cancer death (HR=2.22, 95% CI [1.67, 2.95], *P*=3.2 x10^-8^, **Table 3**). The association of the mortality risk scores calculated by the CpG models with mortality outcomes remained significant after adjusting for age, sex, and clinical risk factors; for all-cause mortality (HR=1.68, 95% CI [1.37, 2.07], *P*=9.8 x10^-7^), CVD death (HR=1.81, 95% CI [1.24, 2.64], *P*=0.002), and cancer death (HR=2.04, 95% CI [1.46, 2.86], *P*=3.0x10^-5^).

#### The integrative model (trained by CpGs and clinical risk factors) moderately improved upon the clinical risk factor model for all-cause mortality and CVD death, and greatly improved the prediction of cancer death

As shown in **Table 3**, the integrative models demonstrated robustness for predicting mortality outcomes, with a good C-index, HR, and low brier error rate. The final integrative models trained using data from all FHS participants are provided in **Table S14** including nine clinical risk factors and 36 CpGs to predict all-cause mortality and CVD death, and in **Table S15** including seven clinical risk factors and 42 CpGs to predict cancer death (excluding prevalent cancer cases). The C-index values of the integrative models were 0.80 (FHS, reflecting the average values of 10-fold cross-validation) and 0.77 (ARIC) for all-cause mortality; 0.83 (FHS) and 0.80 (ARIC) for CVD death; and 0.69 (FHS) and 0.76 (ARIC) for cancer death. Kaplan-Meier survival curves for the mortality risk scores (split into high, middle, and low risk groups) in the ARIC EA cohort (computed by the integrative models using clinical risk factors and CpGs at discovery *P*<1x10^-6^, **Table S14-S15**) illustrates the higher death rate for those with a higher mortality risk score (log-rank *P*<1x10^-6^, **Fig. 3**). In comparison to the clinical risk factor models, the integrative models moderately improved prediction of all-cause mortality (0.7% increase in C-index with addition of CpGs in FHS and 2% increase in ARIC), and of CVD death (2% increase in C-index in FHS, but no increase in ARIC). We speculate that the reason for this minor increase is because the mortality-related CpGs capture the contributions of clinical risk factors for CVD death. For cancer death, the C-index of the integrative model revealed an 11% increase in FHS above and beyond the clinical risk factor model and a corresponding 5% increase in ARIC.

**Figure 3:**
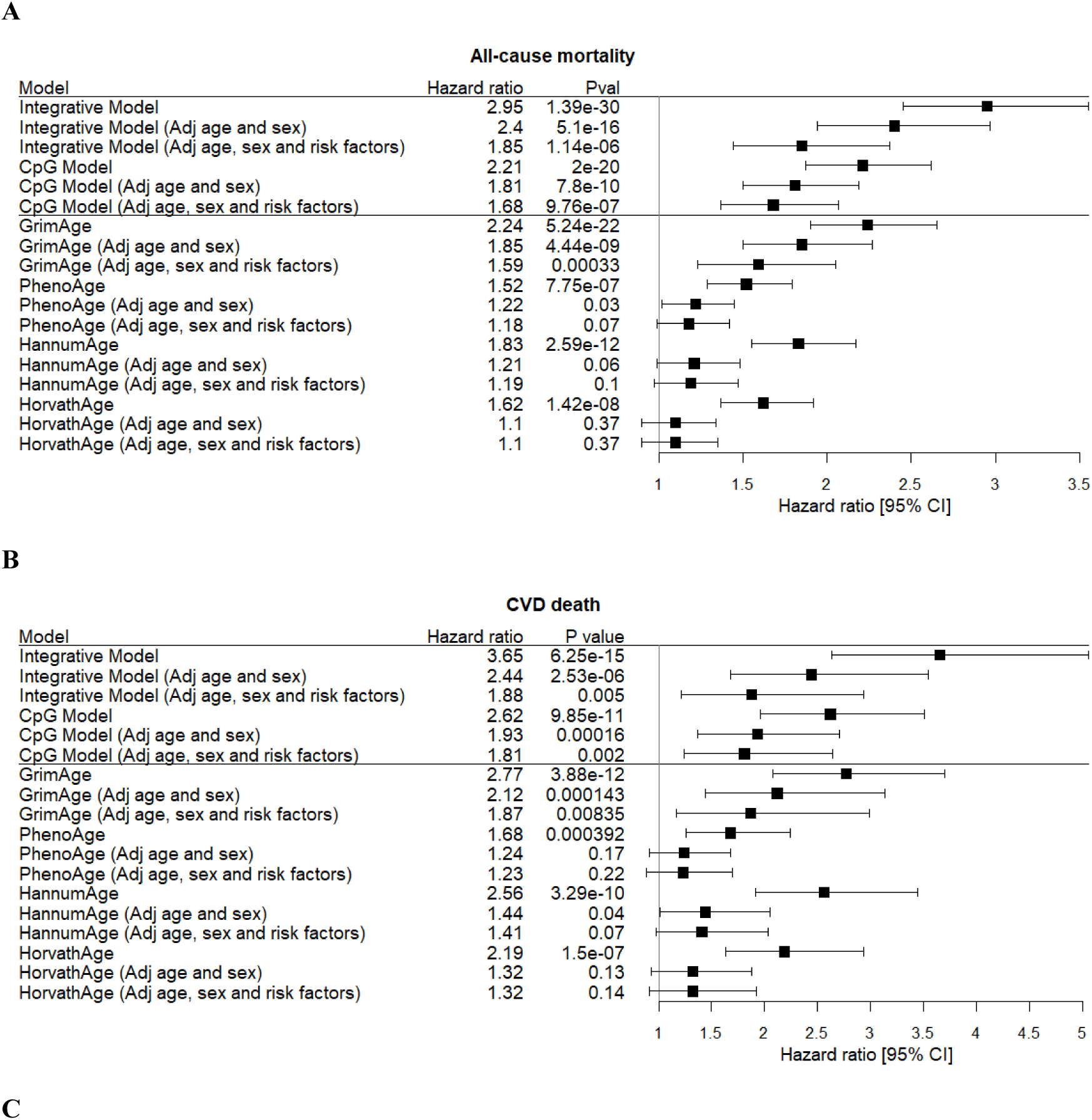
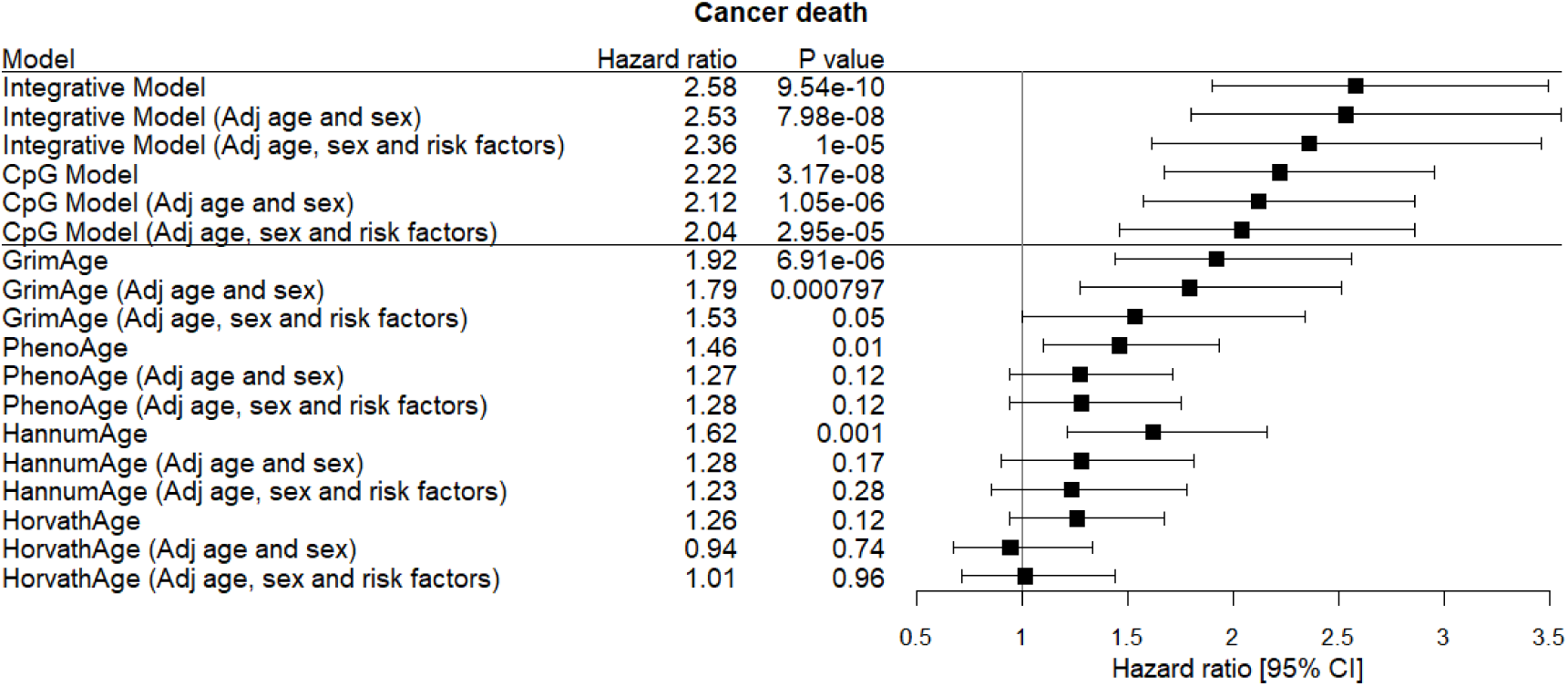
Hazard ratios per standard deviation increment with 95% confidence intervals for mortality. A) with respect to all-cause mortality; B) with respect to CVD death; C) with respect to cancer death. The results were obtained from ARIC European ancestry participants with follow up truncated at 15 years. For cancer death, samples who had any type of cancer before blood drawn for DNA methylation measurements were excluded. Cox regression models were used to relate mortality outcomes to inversely-transformed mortality risk scores computed by Integrative models (**Table S12-S13**) and CpG models (**Table S10-S11**), and inversely-transformed DNAm age including GrimAge (Lu et al., 2019), PhenoAge(Levine et al., 2018), Horvath Age(Horvath, 2013), and Hannum Age(Hannum et al., 2013). *Adj age and sex* indicated the association further adjusted for age and sex. *Adj age, sex and risk factors* indicated the association further adjusted for age, sex and the other clinical risk factors.

We also tested the mortality prediction models’ performance using the entire ARIC EA data (without truncation, **Table S16**). Due to the long follow-up time in this older cohort (mean age 59.8 at baseline, with 20 ± 5.5 years follow-up), the integrative model exhibits very similar performance features as the model using age and sex as the sole input features for predicting all-cause mortality and CVD death. The integrative model improved prediction of cancer death with 2% increase in the C-index versus the clinical risk factor model.

We further tested all-cause mortality prediction models in the CARDIA study (baseline age 45 ± 3 years). The CARDIA study has 12 years of follow-up, during which there were 27 deaths from all causes in 905 participants with DNA methylation. As shown in **Table S17**, the clinical risk factor model, the CpG model, and the integrative model each predicted all-cause mortality, and each outperformed the DNAm age models.

### Comparing the mortality prediction model with DNAm age

We compared four DNAm age models (i.e., PhenoAge (Levine et al., 2018), Horvath Age (Horvath, 2013), Hannum Age (Hannum et al., 2013), and GrimAge (Lu et al., 2019)) with our mortality prediction models (CpG only models and integrative CpG plus 12 risk factor models) for all-cause mortality, CVD death, and cancer death in ARIC participants. The associations of mortality risk scores calculated by mortality prediction models with mortality outcomes were statistically significant, and the associations remained significant after adjusting for age and sex, and after additionally adjusting for the clinical risk factors. The four DNAm age models were significantly associated with mortality outcomes. After adjusting for age, sex and clinical risk factors, however, only GrimAge remained associated with all-cause mortality, CVD death, and cancer death. None of the other three DNAm age predictors was associated with mortality outcomes after additionally adjusting for clinical risk factors (**Fig. 3**). The mortality prediction models (both the CpG only model and the integrative model that included the clinical risk factors and CpGs) outperformed the GrimAge model in prediction of mortality outcomes in terms of HRs and P values. The associations of mortality risk scores with mortality outcomes remain significant after adjusting for the four DNAm age (**Table S18**).

### Associations of DNAm with genetic variants and Mendelian randomization analysis

Among the 177 all-cause mortality-related CpGs (union of EA and AA results at *P*<1x10^-7^), 123 CpGs had significant associations with genetic variants (i.e. *cis-* or *trans-*meQTL variants). meQTL variants for 80 CpGs could be linked to 618 GWAS Catalog(Buniello et al., 2019) index SNPs associated with 432 complex traits or diseases (**Table S18**).

We further performed multiple instrumental variable (IV) MR analysis for the 17 CpGs having ≥ independent *cis-*meQTL SNPs (pruned by LD *r^2^*< 0.01, as IVs, to model the causal relations of differential methylation at these CpGs (as the exposure) on the various outcomes, including longevity (Deelen et al., 2019), CVD, CVD risk factors, and cancer (Evangelou et al., 2018; Locke et al., 2015; Michailidou et al., 2017; Phelan et al., 2017; Schumacher et al., 2018; Scott et al., 2017; Wang et al., 2014; Willer et al., 2013). At *P_MR_* <0.05, MR supported causal effects of 15 CpGs on one or more outcome (**Table S19**), and 4 CpGs were statistically significant at *P_MR_* <0.05/17, including cg06885782 (within 1500 bases upstream of transcription start site [TSS1500] of *KCNQ4*) and cg04907244 (TSS1500 of *SNORD93*) in relation to prostate cancer (Schumacher et al., 2018) (Beta=1.2 and 2.1; and *P_MR_*= 4.1x10^-4^ and 0.003 , respectively), cg07094298 (in the gene body of *TNIP2*) in relation to lung cancer (Wang et al., 2014) (Beta =2.2, and *P_MR_*=0.003), and cg18241337 (in the gene body of *SSR3*) in relation to total cholesterol (Willer et al., 2013) (Beta=0.5, and *P_MR_*=0.003). cg06885782 (*KCNQ4*) also was associated with longevity (Deelen et al., 2019) (Beta=-1.9, *P_MR_*=0.02).

### Associations of DNAm with gene expression, and pathway analysis

For the 177 all-cause mortality-related CpGs at *P*<1x10^-7^, we assessed associations of CpGs with nearby gene expression (i.e. *cis* gene expression; within +/-1 Mb) and identified 15 *cis-* DNAm-mRNA associated pairs (13 CpGs and 15 mRNAs) at *P*<3x10^-10^. The genes located at these CpGs or *cis-*eQTM mRNAs were not enriched for any biological processes or pathways. For the 719 all-cause mortality-related CpGs at *P*<1x10^-5^, 495 genes located at CpG sites were enriched for positive regulation of transcription from RNA polymerase II promoter (Gene Ontology [GO] (Ashburner et al., 2000), fold change = 1.9, FDR=0.05), and pathways for cancer (Kyoto Encyclopedia of Genes and Genomes [KEGG] pathway (Kanehisa & Goto, 2000), fold change =2.2, FDR=0.13, **Table S20**). There were 79 *cis-*DNAm-mRNA pairs (63 CpGs and 67 mRNAs, **Table S21**). The 67 *cis-*eQTM mRNAs were enriched for multiple immune functions including immune response (GO, fold change = 6.3, FDR=0.01).

## Discussion

By performing EWAS using whole blood derived DNA from 15,013 individuals from 15 cohorts with the accrual of 4314 deaths during a mean follow up of more than 10 years, we identified robust DNAm signatures of all-cause and cause-specific mortality. We developed replicable mortality predictors by integrating mortality-related CpGs with traditional clinical risk factors. The integrative models that included clinical risk factors and CpGs showed modest improvement in prediction of all-cause mortality and CVD death, and a substantial improvement in prediction of cancer death compared to the traditional risk factor model in the FHS (internal cross-replication) and ARIC (external independent replication) cohorts.

Our study is one of the largest EWAS of mortality to date (Colicino et al., 2020; Svane et al., 2018; Zhang et al., 2017) and it revealed many replicable DNAm signatures for all-cause mortality. Our results are consistent with those from previous EWAS of all-cause mortality; the vast majority of CpGs (85% in our study, 84% in (Zhang et al., 2017), and 67 % in (Colicino et al., 2020)) were inversely associated with mortality suggesting a greater mortality risk with lower CpG methylation. Our study identified more CpGs in EA cohorts (n=163) than in AA cohorts (n=17). As shown in **Table 2**, the effect sizes (i.e., HR) of mortality-related CpGs in EA and AA participants were quite similar. We speculate that our study identified many more CpGs in EA participants than AA participants due the greater statistical power of the larger EA sample size. Using different DNAm data normalization methods (such as Noob (Triche Jr, Weisenberger, Van Den Berg, Laird, & Siegmund, 2013), SWAN (Maksimovic, Gordon, & Oshlack, 2012), BMIQ(Teschendorff et al., 2013), and Dasen (Pidsley et al., 2013), see **Additional File 1**) in different cohorts may also affect the reproducibility of the results. Among the top mortality-associated CpGs, many were associated with common traits associated with diseases in prior EWAS including BMI (e.g., cg03725309 in *SARS*) (Mendelson et al., 2017), smoking (e.g., cg05575921 in *AHRR*) (Joehanes et al., 2016), blood pressure (e.g., cg03068497 and cg21429551 in *GARS*) (Richard et al., 2017), alcohol consumption (e.g., cg02583484 in *HNRNPA1*) (Liu et al., 2018), and diet (e.g., cg18181703 in *SOCS3* ) (Ma et al., 2020).

Among the 177 all-cause mortality-related CpGs (union of EA and AA results at *P*<1x10^-7^), 123 CpGs had significant associations with genetic variants (i.e., *cis-* or *trans-*meQTL variants identified previously (Huan et al., 2019)). For the remaining 44 CpGs, however, this does not mean that their methylation levels have nothing to do with genetic variation. It is possible that the previous meQTL study lacked sufficient statistical power to identify meQTLs for those CpGs. The mortality-related CpGs are linked to hundreds of human complex diseases/traits via their *cis-*meQTL SNPs, which coincide with 618 GWAS Catalog (Buniello et al., 2019) index SNPs. This leads us to hypothesize that many disease/phenotype associated SNPs may contribute to disease processes via effects on mortality-related CpGs. In this way, the mortality-related CpGs may contribute causally to disease. To test this hypothesis, we conducted MR analyses that confirmed several putatively causal associations of mortality-related CpGs with longevity (Deelen et al., 2019), CVD (Nikpay et al., 2015), CVD risk factors, and several types of cancer (Evangelou et al., 2018; Locke et al., 2015; Michailidou et al., 2017; Phelan et al., 2017; Schumacher et al., 2018; Scott et al., 2017; Wang et al., 2014; Willer et al., 2013) (**Table S19**). Among the four CpGs passing a Bonferroni-corrected threshold in MR analyses, cg06885782 in *KCNQ4* was reported to be associated with risk for prostate cancer (beta=1.2, *P_MR_*=4.1x10^-4^), and negatively associated with longevity (beta=-1.9, *P_MR_*=0.02). *KCNQ4* (potassium voltage-gated channel subfamily Q member 4) was previously reported to be associated with age-related hearing impairment (Van Eyken et al., 2006), and it contains genetic variants associated with all-cause mortality and survival free of major diseases (Walter et al., 2011). cg07094298 in the gene body of *TNIP2* was previously identified as causal for lung cancer. A recent study reported *TNIP2-ALK* fusion in lung adenocarcinoma (Feng et al., 2019). cg04907244 (in TSS1500 of *SNORD93*) was identified as causal for prostate cancer by MR. *SNORD93* and its methylation was reported to be associated with several cancer types including uveal melanoma (Gong et al., 2017), breast cancer (Patterson et al., 2017), and renal clear cell carcinoma (Zhao et al., 2020). Pathway analysis further supported a role of mortality-related CpGs in relation to cancer risk. The intragenic CpGs were enriched for genes in cancer pathways, possibly as a consequence of the expression of nearby genes (*cis-*eQTMs analysis, **Table S21**) related to immune function.

The 14 clinical risk factors for mortality were chosen based on prior knowledge. In contrast, there are far fewer established risk factors for cancer death other than age, sex, BMI, smoking, and alcohol consumption. It is not a surprise that the clinical risk factors themselves accurately predicted all-cause mortality (C-index = 0.80 in FHS, and 0.75 in ARIC) and CVD death (0.81 in FHS and 0.81 in ARIC), but not cancer death (0.57 in FHS and 0.71 in ARIC). Even though the clinical risk factors are important for stratifying CVD risk, clinical risk factors themselves are unable to reveal molecular mechanism and are thereby unable to highlight causal mechanisms or promising therapeutic targets.

After integrating clinical risk factors with DNAm in the all-cause mortality prediction model, the C-index only slightly increased (less than 2%) compared with the clinical risk factors model with regard to all-cause mortality and CVD death. As shown in **Table S14**, nine of the 14 clinical risk factors, including age, sex, physical activity, prevalent cancer, type II diabetes, hypertension, CHD, heart failure and stroke, as well as 36 CpGs that were selected as the representative features. Compared with clinical risk factors, the individual coefficients of the CpGs are much smaller. The small increase in the C-index and the small coefficients of the CpGs suggest that the contribution of CpGs to the prediction of death may overlap with these clinical risk factors. We also found that the mortality-related CpGs as the sole input features were still able to predict mortality outcomes after adjusting for clinical risk factors. This suggests that mortality-related CpGs may identify novel molecular mechanisms contributing to CVD mortality that cannot be captured by existing clinical risk factors.

In contrast to CVD and CVD mortality, for which established risk factors are highly predictive of risk, the prediction of cancer and cancer mortality has proved much more challenging. Owing to the lower prediction using clinical risk factors alone (0.57 in FHS and 0.71 in ARIC), the mortality-related CpGs improved risk prediction of cancer death over and above the clinical risk factor model with an 11% increase in the C-index in FHS and a 5% increase in ARIC. We further tested whether the all-cause mortality prediction model can be used to predict mortality among all participants in the FHS with prevalent cancer (n=389). During a mean follow up of 9 years, there were 165 deaths in this group. The integrative mortality model predicted mortality risk among cancer cases (HR [95%CI]: 4.23 [2.63-6.80], *P* = 2.9x10^-9^). These results in conjunction with MR and pathway analysis, show strong evidence of potential causal relations between mortality-related CpCs and pathways in cancer. Based on these results, we hypothesize that mortality-related CpGs can shed light on the epigenetic regulation of molecular interactions and help to identify novel therapeutic targets to reduce mortality risk for both CVD and cancer death.

Recent studies have used DNAm of multiple CpG sites to predict chronological age (i.e. DNAm age), and showed that DNAm age was associated with all-cause mortality. We explored the prediction provided by these DNAm age models and show that PhenoAge (Levine et al., 2018), Horvath Age (Horvath, 2013), Hannum Age (Hannum et al., 2013), and GrimAge (Lu et al., 2019) were associated with mortality before accounting for risk factors. Only GrimAge, however, remained associated with mortality after adjusting for clinical risk factors. In contrast, the other three DNAm age models were no longer associated with mortality (**Fig. 3**). One possible explanation is that the three DNAm age predictors (i.e., PhenoAge, Horvath Age, and Hannum Age) identify CpGs associated with age, but are not specific for all-cause or cause-specific mortality risk. Of note, the CpGs that serve as DNAm mortality predictors and those that predict DNAm age in the three models do not overlap. Among the top CpGs (N=177) associated with all-cause mortality in our EWAS, only cg00687674 in *TMEM84* is included in PhenoAge (Levine et al., 2018), and only cg19935065 in *DNTT* appears in Hannum Age (Hannum et al., 2013). GrimAge may have outperformed the other three DNAm age models in predicting mortality because the CpGs that it uses are associated with the levels of 80 CVD-related blood proteins, and with lifestyle and clinical risk factors (such as smoking), and mortality (Ho et al., 2018; Shah et al., 2019; Yao et al., 2018). However, because the CpGs in the GrimAge model are not disclosed (i.e. they are proprietary), we were unable to determine if any of the mortality-related CpGs in our study overlap with CpGs in the GrimAge model. Of note, our mortality prediction models (both the CpG only model and the integrative model that included CpGs and the clinical risk factors) outperformed GrimAge in prediction of mortality outcomes.

We tested and compared four prediction methods including Elastic-coxph (Friedman et al., 2010), a regression based method, and three machine learning methods (Ching et al., 2018; Ishwaran et al., 2008; Katzman et al., 2018). The machine learning models did not outperform Elastic-coxph (**Table S9** and **Fig. S3**). The clinical risk factor model trained by machine learning methods did not perform well in independent external replication. For example, the C-index of the clinical risk factor model for all-cause mortality was 0.67 using RSF^17^ versus 0.75 using Elastic-coxph in ARIC participants. Based on this metric, the machine learning methods did not outperform the regression-based methods when there were relatively few features as inputs.

The primary outcome of our study was all-cause mortality. We did not train prediction models for CVD death or cancer death, but we tested the prediction ability of the all-cause mortality predictor on CVD death and cancer death. The CpGs in the model were restricted to all-cause mortality related CpGs. As shown in **Fig. 1**, the top DNAm signatures for all-cause mortality showed the same direction of effect for CVD death and cancer death. It is possible that some CpGs show opposite directions in relation to CVD death and cancer death, but we did not train separate models for these outcomes. Therefore, developing separate prediction models for CVD death and cancer death with a very large sample size would be an important next step.

## Conclusions

In conclusion, the ancestry-stratified epigenome-wide meta-analyses in 15 population-based cohorts identified replicable DNAm signatures of all-cause and cause-specific mortality. The top mortality-associated CpGs were linked with genes involved in immune and cancer related pathways, and were reported to be associated with human longevity, CVD risk factors, and several types of cancer. We constructed and validated DNAm-based prediction models that predicted mortality risk independent of established clinical risk factors. The prediction model trained by integrating DNAm with clinical risk factors showed modest improvement in prediction of all-cause mortality and CVD death, and a substantial improvement in prediction of cancer death, compared with the model trained by clinical risk factors alone. The mortality-related CpG sites, and the DNAm-based prediction models may serve as useful clinical tools for assessing all-cause and cause-specific mortality risk and for developing new therapeutic strategies.

## Methods

### Study population

This study included 15,013 participants from 15 population-based cohorts. There were 11,684 European ancestry (EA) participants from 12 cohorts, including the Atherosclerosis Risk in Communities (ARIC) Study, the Cardiovascular Health Study (CHS), the Danish Twin Register sample (DTR), the Epidemiologische Studie zu Chancen der Verhütung, Früherkennung und optimierten Therapie chronischer Erkrankungen in der älteren Bevölkerung (ESTHER), the Framingham Heart Study (FHS), the Invecchiare in Chianti (InCHIANTI) Study, the Cooperative Health Research in the Region of Augsburg (KORA F4), the Lothian Birth Cohorts of 1921 (LBC1921) and 1936 (LBC1936), the Normative Aging Study (NAS), the Rotterdam Study (RS), and Women’s Health Initiative (WHI); and 3329 Africa ancestry (AA) participants from 3 cohorts, including ARIC, CHS, and WHI. For each participant, we calculated the follow-up time between the date of the blood draw for DNAm measurements and the date at death or last follow up. Mean follow up was less than 15 years (range 6.2 to 13.7) for most cohorts, except for ARIC (mean 20.0 for EA and 18.6 for AA). The protocol for each study was approved by the institutional review board of each cohort. Further details for each cohort were included in **Additional file1**.

### Mortality ascertainment and clinical phenotypes

Outcomes including death from all causes, deaths from CVD, and deaths from cancer, were prospectively ascertained in each cohort. Survival status and details of death were ascertained using multiple strategies, including routine contact with participants for health history updates, surveillance at the local hospital, review of obituaries in the local newspaper, and National Death Index queries. Death certificates, hospital and nursing home records prior to death, and autopsy reports were requested and reviewed. Date and cause of death were determined separately for each cohort following review of all available medical records and /or were register-based.

The clinical and lifestyle risk factors (referred to as clinical risk factors for simplicity thereafter) used as covariates in this study included age, sex, body mass index (BMI), smoking, alcohol consumption, physical activity, educational attainment, and prevalent diseases including hypertension, coronary heart disease (CHD), heart failure, stroke, type-II diabetes, and cancer. Fourteen clinical risk factors were chosen based on prior knowledge; most of these are key CVD risk factors. The clinical risk factors were ascertained at the time of blood draw for DNAm measurements. BMI was calculated as weight (kg) divided by height squared (m^2^). Educational attainment (years of educational schooling), physical activity (frequency, intensity or the metabolic equivalent of task [MET] scores), smoking status (yes/no, or cigs/day), alcohol consumption (drinks per day) were self-reported or ascertained by an administered questionnaire at routine research clinic visits. Diabetes was defined as a measured fasting blood glucose level of >125 mg/dL or current use of glucose-lowering prescription medication. Hypertension was defined as a measured systolic blood pressure (BP) ≥ 140 mm Hg or diastolic BP ≥ 90 mm Hg or use of antihypertensive prescription medication. Cancer was defined as the occurrence of any type of cancer excluding non-melanoma skin cancer.

### DNA methylation measurements and quality control

For each cohort, DNA was extracted from whole blood and bisulfite-converted using a Zymo EZ DNA methylation kit. DNAm was measured using the Illumina Infinium HumanMethylation450 (450K) BeadChip platform (Illumina Inc, San Diego, CA). Each cohort conducted independent laboratory DNAm measurement, quality control (including sample-wise and probe-wise filtering, and probe intensity background correction; see **Additional file1**).

### Cohort-specific epigenome-wide association analysis

The correction of methylation data for technical covariates was cohort specific. Each cohort performed an independent investigation to select an optimized set of technical covariates (e.g., batch, plate, chip, row and column), using measured or imputed blood cell type fractions, surrogate variables, and/or principal components. Most cohorts had previous publications using the same dataset for EWAS of different traits, such as EWAS of alcohol drinking and smoking (Mendelson et al., 2017; Michailidou et al., 2017). In this study, those cohorts used the same strategies as they did previously for correcting for technical variables including batch (see **Additional file1**). To avoid false positives driven by single CpG extreme values, in each cohort, we first performed rank-based inverse normal transformation (INT) of DNAm β-values (the ratio of methylated probe intensity divided by the sum of the methylation and unmethylated probe intensity). We then conducted time-to-event analyses using Cox proportional hazards models to test for associations between each CpG and mortality outcomes including all-cause mortality, CVD death, and cancer death using the *coxph*() function in the ‘survival’ R library, adjusting for clinical risk factors (see **Mortality ascertainment and clinical risk factors**), technical confounders, and familial relatedness. Because ARIC cohorts had much longer follow-up than the other cohorts, ARIC follow up was truncated at 15 years and results were compared to those before truncation to determine if results were impacted by duration of follow up.

In this study, we performed INT of DNAm β-values to avoid false positives driven by extreme values of single CpGs. Using the FHS EWAS results as an example, **Table S21** shows that the top CpGs associated with all-cause mortality (without INT) were no longer significant after performing INT. This finding suggests that if we directly use DNAm β-values, those extreme outlier values could lead to false positive results. Clearly, the distribution of DNA β-values is non-normal and for this reason, we believe that the conservative INT approach we took protected against false positive results.

### Meta-analysis

The meta-analysis was performed for all-cause mortality, CVD death, and cancer death in EA (n=11,684) and AA (n=3329) participants respectively, using inverse variance-weighted random-effects models implemented in *metagen()* function R packages (https://rdrr.io/cran/meta/man/metagen.html). We chose a random-effects model because of the heterogeneity in follow-up length and population demographics in the different cohorts (**Table S1**). We excluded the EWAS results for a study with <20 deaths. We excluded probes mapping to multiple locations on the sex chromosomes or with an underlying SNP (MAF>5% in 1000 Genome Project data) at the CpG site or within 10bp of the single base extension. In addition, the meta-analysis was constrained to methylation probes passing filtering criteria in five or more cohorts (see **Additional File1**), which resulted in ∼400,000 CpGs that were included in the final analyses. The statistical significance threshold was *P*<0.05/400,000 ≈ 1x10^-7^.

Three types of sensitivity analyses were performed including 1) correcting for λ values in each cohorts (Devlin, Roeder, & Wasserman, 2001), 2) excluding two cohorts with λ >1.5 from the meta-analysis, and 3) excluding results of RS, because the cohort-specific analysis in RS having a strange distribution of top hits. There were 157 CpGs identified at *P*<1e-7 in the RS cohort-specific analysis. The number is much more than the number of all-cause mortality associated CpGs identified in the other cohorts.

### Mortality prediction models

Mortality prediction models based on clinical risk factors and with the addition of DNAm were built and tested in EA cohorts. The analysis flowchart is shown in **Fig. S1**. To ensure unbiased validation, we split the EA cohorts into discovery and replication sets. The discovery cohorts consisted of 8288 participants from 10 cohorts, excluding FHS (n=2427) and ARIC (n=969), which were used as replication cohorts. To build and replicate a prediction model, the DNAm data were preprocessed utilizing the same strategy as in the EWAS analysis.

#### Input features

To evaluate the prediction performance of clinical risk factors and DNAm comprehensively, we tested 13 sets of features, Feature set 1 (**F1**) included age (years), sex (male as 1 and female as 2), and 12 other clinical risk factors including BMI (kg/m2), smoking (current smoker as 1, and former and never smoker as 0), alcohol consumption (grams/day), physical activity (MET scores), educational attainment (education years), and prevalent diseases (yes as 1 and no as 0) including hypertension, CHD, heart failure, stroke, type-2 diabetes, and cancer. **F2-F7** were mortality-related CpGs selected by meta-analysis in the discovery cohorts by inverse-variance weighted random-effects models at a series of p value thresholds, including **F2** CpGs at *P*<1e-7, **F3** CpGs at *P*<1e-6, **F4** CpGs at *P*<1e-5, **F5** CpGs at *P*<1e-4, **F6** CpGs at *P*<1e-3, and **F7** CpGs at *P*<0.05. **F8-F13** are **F1** (age, sex and 12 clinical phenotypes) plus **F2-F7** respectively. In doing so, we were able to evaluate the prediction performance based on the clinical risk factors (**F1**) and the DNAm (**F2-F7**), and test if the combination of DNAm with clinical risk factors (**F8-F13**) could be able to improve the prediction performance by using clinical risk factors (**F1**) only and DNAm only (**F2-F7**).

#### Model building

We compared four methods of building prediction models, including 1) Elastic net -Cox proportional hazards method (Elastic-coxph, using *glmnet*, a R package) (Friedman et al., 2010); 2) Random survival forest (RSF, using *randomForestSRC*, a R package) (Ishwaran et al., 2008); 3) Cox-nnet (https://github.com/lanagarmire/cox-nnet, a Python package) (Ching et al., 2018), and 4) DeepSurv (https://github.com/jaredleekatzman/DeepSurv, a Python package) (Katzman et al., 2018). The first method is a penalized linear regression method, while the other three are non-linear machine learning methods.

*Elastic-coxph* is a Cox regression model regularized with elastic net penalty (Friedman et al., 2010). Performing this method requires to identify best values of two parameters, α and λ. We tuned each model by iterating over a number of α and λ values under cross-validation. α indicated linearly combined penalties of the lasso (α=0) and ridge (α=1) regression. λ is the shrinkage parameter, when λ =0 indicated no shrinkage, and as λ increases, the coefficients are shrunk ever more strongly. Effectively this will shrink some coefficients close to 0 for optimizing a set of features. The α value was set to 0.5, and the λ value was set to lambda.min when training models.

*RSF* is an ensemble tree model that is based on the random forest method for survival analysis (Ishwaran et al., 2008). The optimized values of parameters in RSF models, including the number of trees (nTrees=100) and nodeSize =15 were chosen by iterating over a number of values which maximized the accuracy of RSF models tested in the replication sets under cross-validation. RSF can compute feature importance scores for feature selection.

*Cox-nnet* is an artificial neural network based method for survival analysis (Ching et al., 2018). Cox-nnet includes two layer neural network: one hidden layer and one output layer. The output layer was used to perform Cox regression based on the activation levels of the hidden layer. Cox-nnet could also compute feature importance scores for feature selection. For each model training, the *L2* regularization parameter is optimized using the *L2CVProfile* Python function by iterating over a number of values under cross-validation.

*DeepSurv* is a deep learning-based survival prediction method (Katzman et al., 2018). DeepSurv uses a multi-layer feed forward neural network, of which the hidden layers consist of a fully-connected layer of nodes, followed by a dropout layer, and the output is a single node with a linear activation which estimated the log-risk function in the Cox model, parameterized by the weight of the network. The values of hyperparameters when using DeepSurv were *L2* regularization = 0.8, dropout = 0.4, learning rate = 0.02, hidden layer size (4 layers with nodes 500, 200, 100 and 50), lr_decay = 0.001, momentum = 0.9 and the activation method (using Scaled Exponential Linear Units), which were optimized by iterating over a number of values each-by-each and under cross-validation. *DeepSurv* has not been used previously for selecting features.

#### Cross-validation

The 2427 FHS participants were randomly split into 5 equal sets (n=485 or 486 in each set), and each set included approximately equal numbers of deaths. We then used 3 of the 5 sets (60%) for model training and the remaining 2 sets (40%) for model testing. In doing so, we obtained 10 combinations. In each training / testing combination, we constructed a model using the training data, and then used the model to generate a mortality risk score based on the testing data. We assessed associations of the predicted mortality risk score (after inverse normal transformation) with all-cause mortality, CVD death, and cancer death in the testing data using time-to-event proportional hazards models. This data partitioning and cross-validation strategy was only used to assess the robustness of prediction models when using different features and methods, and to select the optimized parameters for training models. The final models reported were built on all FHS participants using the optimized parameters. We also repeated the same analysis steps using FHS participants without cancer at baseline (n=2038; 238 deaths from all causes, 70 from CVD, and 42 from cancer).

#### Independent external validation

The prediction models built using all FHS participants were further tested in ARIC EA participants for the prediction of mortality outcomes. We performed tests on all-cause mortality and CVD death on all ARIC EA participants truncated at 15 years of follow up, and tests on cancer death after excluding prevalent cancer.

#### Evaluation of model performance

We used four evaluation metrics to assess model performance, including the concordance index (C-index) (Harrell Jr, Lee, & Mark, 1996), hazards ratio of predicted risk score (inversely-transformed) for prediction of mortality, the integrated brier score (IBS) (Brier, 1950), and Kaplan-Meier (KM) survival curves for high, medium and low risk groups (Kaplan & Meier, 1958). The C-index reflects the percentage of individuals whose predicted survival times are correctly ordered. A C-index of 0.50 reflects no improvement in prediction over chance. The brier score measures the mean of the difference between the observed and the estimated survival beyond a certain time. The brier score ranges between 0 and 1, and a larger score indicates higher inaccuracy. The integrated brier score is the brier score averaged over the entire time interval.

### DNAm Age

We compared the prediction performance of DNAm age with our DNAm-based mortality prediction model in relation to all-cause mortality, CVD death, and cancer death in the ARIC EA cohort (truncating follow up at 15 years). Four measures of DNAm age were used in this study, including PhenoAge (Levine et al., 2018), Horvath age (Horvath, 2013), Hannum age (Hannum et al., 2013) and GrimAge (Lu et al., 2019). The Horvath Age is based on 353 CpGs, the Hannum age is based on 71 CpGs, and PhenoAge is based on 513 CpGs. DNAm age was calculated as the sum of the beta values multiplied by the reported effect size. Due to the GrimAge model was not publicly available, the GrimAge was calculated by uploading the DNAm data to the website (http://dnamage.genetics.ucla.edu/). Proportional hazards regression models were used to test the association between inversely-rank transformed DNAm age (all 3 approaches) and mortality outcomes, adjusting for age, sex, and clinical covariates (see **Mortality ascertainment and clinical phenotypes**).

### meQTLs

meQTLs (SNPs associated with DNA methylation) were identified from 4170 FHS participants as reported previously, including 4.7 million *cis-*meQTLs and 630K *trans-*meQTLs at *P*<2x10^-11^ for *cis* and *P*<1.5x10^-14^ for *trans* (Huan et al., 2019). The genotypes were measured using Affymetrix SNP 500K mapping and Affymetrix 50K gene-focused MIP arrays. Genotypes were imputed using the 1000 Genomes Project panel phase 3 using MACH / Minimac software. SNPs with MAF >0.01 and imputation quality ratio >0.3 were retained. *cis-*meQTLs were defined as SNPs residing within 1 Mb upstream or downstream of a CpG site. The FHS meQTL data resource includes 3.5 times more *cis-*, and 10 times more *trans*-meQTL SNPs than the other published studies to date (https://ftp.ncbi.nlm.nih.gov/eqtl/original_submissions/FHS_meQTLs/).

### Mendelian randomization

Two-sample Mendelian randomization (MR) was used to identify putatively causal CpGs for human longevity, CVD and CVD risk factors, and cancer types using a multi-step strategy. Estimated associations and effect sizes between SNPs and traits were based on the latest published GWAS meta-analysis of human longevity (Deelen et al., 2019), coronary heart disease (CHD) (Nikpay et al., 2015); myocardial infarction (MI) (Nikpay et al., 2015); type-II diabetes (T2D) (Scott et al., 2017); body mass index (BMI) (Locke et al., 2015); lipids traits including high-density lipoprotein (HDL) cholesterol, low-density lipoprotein (LDL) cholesterol, total cholesterol (TC), and triglycerides (TG) (Willer et al., 2013); systolic blood pressure (SBP) and diastolic blood pressure (DBP) (Evangelou et al., 2018), and cancer types including breast cancer (Michailidou et al., 2017), prostate cancer (Schumacher et al., 2018), lung cancer (Wang et al., 2014) and ovarian cancer (Phelan et al., 2017). We were unable to include some other popular cancer types, because their GWAS data were not be accessible by us.

Instrumental variables (IVs) for each CpG site consisted of independent *cis-*meQTLs pruned at linkage disequilibrium (LD) *r^2^*<0.01, retaining only one *cis-*meQTL variant with the lowest SNP-CpG *P* value in each LD block. LD proxies were defined using 1000 genomes imputation in EA. Inverse variance weighted (IVW) MR tests were performed on CpGs with at least three independent *cis-*meQTL variants, which is the minimum number of IVs needed to perform multiple instruments MR. The multiple instruments improved the precision of IV estimates, and allowed the examination of underlying IV assumption (Palmer et al., 2012). Among 177 all-cause mortality related CpGs at *P*<1x10^-7^, MR tests were performed on 17 CpGs having ≥3 independent *cis-*meQTL SNPs. To test the validity of IVW-MR results, we performed heterogeneity and MR-EGGER pleiotropy tests for all IVs. The statistical significance threshold for MR is *P_MR_*<0.05/17, and both *P_heter_* and *P_pleio_* were required to be >0.05.

### eQTMs

Association tests of DNAm and gene expression were performed in 3684 FHS participants with available DNAm and gene expression data. mRNA was extracted from whole blood (collected in PAXgene tubes) and profiled using the Affymetrix Human Exon 1.0 ST GeneChip platform. Raw gene expression data were first normalized using the RMA (robust multi-array average) from Affymetrix Power Tools (APT, thermofisher.com/us/en/home/life-science/microarray-analysis/affymetrix.html#1_2) with quantile normalization. Then, output expression values of 17,318 genes were extracted by APT based on NetAffx annotation version 31.

DNAm β values were adjusted for age, sex, predicted blood cell fraction, the two top PCs of DNAm, and 25 surrogate variables (SVs), with DNAm as a fixed effect, and batch as a random effect by fitting LME models. Residuals (DNAm_resid) were retained. The gene expression values were adjusted for age, sex, predicted blood cell fraction, a set of technical covariates, the two top PCs and 25 SVs, with gene expression as a fixed effect, and batch as a random effect by LME, and residuals (mRNA_resid) were retained. Then, linear regression models were used to assess pair-wise associations between DNAm_resid and mRNA_resid. SVs were calculated using the SVA package in R. A *cis-*CpG-mRNA pair was defined as a CpG residing ±1Mb of the TSS of the corresponding gene encoding the mRNA (*cis-*eQTM). The annotations of CpGs and transcripts were obtained from annotation files of the HumanMethylation450K BeadChip and the Affymetrix exon array S1.0 platforms. We estimated that there were 1.6 x 10^8^ potential *cis-* CpG-mRNA pairs. We only used *cis*-eQTMs in this study because *trans-*eQTMs were not replicated in independent external studies. The statistical significance threshold was *P*<3x10^-10^ (0.05 /1.6x10^8^)

### Gene ontology and pathway enrichment analysis

Gene ontology and pathway enrichment analyses were performed on the genes annotated in relation to the 177 all-cause mortality related CpGs at *P*<1x10^-7^or *P*<1x10^-5^ as well as the *cis-*eQTM genes associated with those CpGs. Hypergeometric tests were used to investigate over-representations of genes from multiple biological process and pathways. To improve focus in this study, we only used results of KEGG and Gene Ontology – biological process (GO-BP) terms. Enrichment tests used the online DAVID Bioinformatics Resources 6.8 (https://david.ncifcrf.gov/). The P-value was further corrected by the number of unique GO-BP terms and pathways. A Benjamini-Hochberg corrected FDR<0.2 was considered significant.

### Data Availability

The DNA methylation data and phenotype data are available in dbGaP for some of the cohorts in this study (https://www.ncbi.nlm.nih.gov/gap/) including FHS (accession number phs000724.v5.p10) and WHI (accession number phs000200.v12.p3). For LBC, data are available in the European Genome-phenome Archive (https://www.ebi.ac.uk/ega/home), under accession number EGAS00001000910. For the other cohorts including ARIC, CHS, NAS, InCHIANTi, KORA, ESTHER, Danish, RS and CARDIA, the data are available on request by contacting with the principal investigators of each cohort.

## Declarations

### Ethics approval and consent to participate

This study included participants from 12 population-based cohorts studies, including the Atherosclerosis Risk in Communities (ARIC) Study, the Cardiovascular Health Study (CHS), the Danish Twin Register sample (DTR), the Epidemiologische Studie zu Chancen der Verhütung, Früherkennung und optimierten Therapie chronischer Erkrankungen in der älteren Bevölkerung (ESTHER), the Framingham Heart Study (FHS), the Invecchiare in Chianti (InCHIANTI) Study, the Cooperative Health Research in the Region of Augsburg (KORA F4), the Lothian Birth Cohorts of 1921 (LBC1921) and 1936 (LBC1936), the Normative Aging Study (NAS), the Rotterdam Study (RS), and Women’s Health Initiative (WHI). All of the 12 studies were approved by their institutional review committees (see details in **Additional file 1**). All study participants provided written informed consent.

## Data Availability

https://www.ebi.ac.uk/ega/home

## Acknowledgements

The views expressed in this manuscript are those of the authors and do not necessarily represent the views of the National Heart, Lung, and Blood Institute; the National Institutes of Health; or the U.S. Department of Health and Human Services.

For a list of all the investigators who have contributed to WHI science, please visit: https://s3-us-west-2.amazonaws.com/www-whi-org/wp-content/uploads/WHI-Investigator-Long-List.pdf.

## Funding

The **Framingham Heart Study** is funded by National Institutes of Health contract N01-HC-25195 and HHSN268201500001I. The laboratory work for this investigation was funded by the Division of Intramural Research, National Heart, Lung, and Blood Institute, National Institutes of Health. The analytical component of this project was funded by the Division of Intramural Research, National Heart, Lung, and Blood Institute, and the Center for Information Technology, National Institutes of Health, Bethesda, MD.

The **Cardiovascular Health Study** is supported by NHLBI contracts HHSN268201200036C, HHSN268200800007C, HHSN268201800001C, N01HC55222, N01HC85079, N01HC85080, N01HC85081, N01HC85082, N01HC85083, N01HC85086; and NHLBI grants U01HL080295, U01HL130114, K08HL116640, R01HL087652, R01HL092111, R01HL103612, R01HL105756, R01HL103612, R01HL111089, R01HL116747 and R01HL120393 with additional contribution from the National Institute of Neurological Disorders and Stroke (NINDS). Additional support was provided through R01AG023629 from the National Institute on Aging (NIA), Merck Foundation / Society of Epidemiologic Research as well as Laughlin Family, Alpha Phi Foundation, and Locke Charitable Foundation. A full list of principal CHS investigators and institutions can be found at CHS-NHLBI.org. The provision of genotyping data was supported in part by the National Center for Advancing Translational Sciences, CTSI grant UL1TR000124, and the National Institute of Diabetes and Digestive and Kidney Disease Diabetes Research Center (DRC) grant DK063491 to the Southern California Diabetes Endocrinology Research Center. Infrastructure for the CHARGE Consortium is supported in part by the National Heart, Lung, and Blood Institute grant R01HL105756.

The **DTR** study was supported by The Danish Council for Independent Research—Medical Sciences (DFF-6110-00016), the European Union’s Seventh Framework Programme (FP7/2007–2011) under grant Agreement No. 259679 and The Danish National Program for Research Infrastructure 2007 (09-063256).

The **WHI** program is funded by the National Heart, Lung, and Blood Institute, National Institutes of Health, U.S. Department of Health and Human Services through contracts HHSN268201600018C, HHSN268201600001C, HHSN268201600002C, HHSN268201600003C, and HHSN268201600004C. Work in WHI was NIEHS-supported by R01-ES020836 (EAW; AB; LH).

Phenotype collection in the **Lothian Birth Cohort 1921** was supported by the UK’s Biotechnology and Biological Sciences Research Council (BBSRC), The Royal Society and The Chief Scientist Office of the Scottish Government. Phenotype collection in the **Lothian Birth Cohort 1936** was supported by Age UK (The Disconnected Mind project). Methylation typing was supported by Centre for Cognitive Ageing and Cognitive Epidemiology (Pilot Fund award), Age UK, The Wellcome Trust Institutional Strategic Support Fund, The University of Edinburgh, and The University of Queensland. IJD is a member of the University of Edinburgh Centre for Cognitive Ageing and Cognitive Epidemiology (CCACE), which is supported by funding from the BBSRC, the Medical Research Council (MRC), and the University of Edinburgh as part of the cross-council Lifelong Health and Wellbeing initiative (MR/K026992/1). W.D.H. is supported by a grant from Age UK (Disconnected Mind Project).

## Authors’ contributions

T. H., D.L., and J.P. designed, directed, and supervised the project. T. H., and D.L. drafted the manuscript. T.H., S.N., E.C., C. R., D.H., J.B., M.S., Y.Z., A.B., E.M., and T.T. conducted the analyses. All authors participated in revising and editing the manuscripts. All authors have read and approved the final version of the manuscript.

## Competing Interests

The authors declare no conflict of interest.

## Notes

### Competing Interest Statement

The authors have declared no competing interest.

### Author Declarations

This study included participants from 12 population-based cohorts studies including the Atherosclerosis Risk in Communities ARIC Study the Cardiovascular Health Study CHS the Danish Twin Register sample DTR the Epidemiologische Studie zu Chancen der Verhütung Früherkennung und optimierten Therapie chronischer Erkrankungen in der älteren Bevölkerung ESTHER the Framingham Heart Study FHS the Invecchiare in Chianti InCHIANTI Study the Cooperative Health Research in the Region of Augsburg KORA F4 the Lothian Birth Cohorts of 1921 LBC1921 and 1936 LBC1936 the Normative Aging Study NAS the Rotterdam Study RS and Womens Health Initiative WHI. All of the 12 studies were approved by their institutional review committees see details in Additional file 1. All study participants provided written informed consent.

## References

Ashburner, M., Ball, C. A., Blake, J. A., Botstein, D., Butler, H., Cherry, J. M., . . . Eppig, J. T. (2000). Gene ontology: tool for the unification of biology. Nature genetics, 25(1), 25–29.

Brier, G. W. (1950). Verification of forecasts expressed in terms of probability. Monthly weather review, 78(1), 1–3.

Buniello, A., MacArthur, J. A. L., Cerezo, M., Harris, L. W., Hayhurst, J., Malangone, C., . . . Sollis, E. (2019). The NHGRI-EBI GWAS Catalog of published genome-wide association studies, targeted arrays and summary statistics 2019. Nucleic acids research, 47(D1), D1005–D1012.

Chen, B. H., Marioni, R. E., Colicino, E., Peters, M. J., Ward-Caviness, C. K., Tsai, P.-C., . . . Guan, W. (2016). DNA methylation-based measures of biological age: meta-analysis predicting time to death. Aging (Albany NY), 8(9), 1844.

Ching, T., Zhu, X., & Garmire, L. X. (2018). Cox-nnet: an artificial neural network method for prognosis prediction of high-throughput omics data. PLoS computational biology, 14(4), e1006076.

Colicino, E., Marioni, R., Ward-Caviness, C., Gondalia, R., Guan, W., Chen, B., . . . Golareh, A. (2020). Blood DNA methylation sites predict death risk in a longitudinal study of 12,300 individuals. Aging (Albany NY), 12(14), 14092.

Deelen, J., Evans, D. S., Arking, D. E., Tesi, N., Nygaard, M., Liu, X., . . . Atzmon, G. (2019). A meta-analysis of genome-wide association studies identifies multiple longevity genes. Nature Communications, 10(1), 1–14.

Devlin, B., Roeder, K., & Wasserman, L. (2001). Genomic control, a new approach to genetic-based association studies. Theoretical population biology, 60(3), 155–166.

Dugué, P. A., Bassett, J. K., Joo, J. E., Jung, C. H., Ming Wong, E., Moreno-Betancur, M., . . . Severi, G. (2018). DNA methylation-based biological aging and cancer risk and survival: Pooled analysis of seven prospective studies. International journal of cancer, 142(8), 1611–1619.

Evangelou, E., Warren, H. R., Mosen-Ansorena, D., Mifsud, B., Pazoki, R., Gao, H., . . . Karaman, I. (2018). Genetic analysis of over 1 million people identifies 535 new loci associated with blood pressure traits. Nature genetics, 50(10), 1412–1425.

Feng, T., Chen, Z., Gu, J., Wang, Y., Zhang, J., & Min, L. (2019). The clinical responses of TNIP2-ALK fusion variants to crizotinib in ALK-rearranged lung adenocarcinoma. Lung Cancer, 137, 19–22.

Friedman, J., Hastie, T., & Tibshirani, R. (2010). Regularization paths for generalized linear models via coordinate descent. Journal of statistical software, 33(1), 1.

Gong, J., Li, Y., Liu, C. -j., Xiang, Y., Li, C., Ye, Y., . . . Diao, L. (2017). A pan-cancer analysis of the expression and clinical relevance of small nucleolar RNAs in human cancer. Cell reports, 21(7), 1968–1981.

Hannum, G., Guinney, J., Zhao, L., Zhang, L., Hughes, G., Sadda, S., . . . Gao, Y. (2013). Genome-wide methylation profiles reveal quantitative views of human aging rates. Molecular cell, 49(2), 359–367.

Harrell Jr, F. E., Lee, K. L., & Mark, D.B. (1996). Multivariable prognostic models: issues in developing models, evaluating assumptions and adequacy, and measuring and reducing errors. Statistics in medicine, 15(4), 361–387.

Ho, J. E., Lyass, A., Courchesne, P., Chen, G., Liu, C., Yin, X., . . . Levy, D. (2018). Protein biomarkers of cardiovascular disease and mortality in the community. Journal of the American Heart Association, 7(14), e008108.

Horvath, S. (2013). DNA methylation age of human tissues and cell types. Genome biology, 14(10), 3156.

Horvath, S., Pirazzini, C., Bacalini, M. G., Gentilini, D., Di Blasio, A. M., Delledonne, M., . . . Passarino, G. (2015). Decreased epigenetic age of PBMCs from Italian semi-supercentenarians and their offspring. Aging (Albany NY), 7(12), 1159.

Huan, T., Joehanes, R., Song, C., Peng, F., Guo, Y., Mendelson, M., . . . Richard, M. (2019). Genome-wide identification of DNA methylation QTLs in whole blood highlights pathways for cardiovascular disease. Nature Communications, 10(1), 1–14.

Ishwaran, H., Kogalur, U. B., Blackstone, E. H., & Lauer, M. S. (2008). Random survival forests. The annals of applied statistics, 2(3), 841–860.

Joehanes, R., Just, A. C., Marioni, R. E., Pilling, L. C., Reynolds, L. M., Mandaviya, P. R., . . . Aslibekyan, S. (2016). Epigenetic signatures of cigarette smoking. Circulation: Cardiovascular Genetics, 9(5), 436–447.

Jones, P. A., & Takai, D. (2001). The role of DNA methylation in mammalian epigenetics. Science, 293(5532), 1068–1070.

Kanehisa, M., & Goto, S. (2000). KEGG: kyoto encyclopedia of genes and genomes. Nucleic acids research, 28(1), 27–30.

Kaplan, E. L., & Meier, P. (1958). Nonparametric estimation from incomplete observations. Journal of the American statistical association, 53(282), 457–481.

Katzman, J. L., Shaham, U., Cloninger, A., Bates, J., Jiang, T., & Kluger, Y. (2018). DeepSurv: personalized treatment recommender system using a Cox proportional hazards deep neural network. BMC medical research methodology, 18(1), 24.

Levine, M. E., Lu, A. T., Quach, A., Chen, B. H., Assimes, T. L., Bandinelli, S., . . . Li, Y. (2018). An epigenetic biomarker of aging for lifespan and healthspan. Aging (Albany NY), 10(4), 573.

Liu, C., Marioni, R. E., Hedman, Å. K., Pfeiffer, L., Tsai, P.-C., Reynolds, L. M., . . . Tanaka, T. (2018). A DNA methylation biomarker of alcohol consumption. Molecular psychiatry, 23(2), 422–433.

Locke, A. E., Kahali, B., Berndt, S. I., Justice, A. E., Pers, T. H., Day, F. R., . . . Yang, J. (2015). Genetic studies of body mass index yield new insights for obesity biology. Nature, 518(7538), 197–206.

Lu, A. T., Quach, A., Wilson, J. G., Reiner, A. P., Aviv, A., Raj, K., . . . Stewart, J. D. (2019). DNA methylation GrimAge strongly predicts lifespan and healthspan. Aging (Albany NY), 11(2), 303.

Ma, J., Rebholz, C. M., Braun, K. V., Reynolds, L. M., Aslibekyan, S., Xia, R., . . . Mendelson, M. M. (2020). Whole Blood DNA Methylation Signatures of Diet Are Associated with Cardiovascular Disease Risk Factors and All-cause Mortality. Circulation: Genomic and Precision Medicine.

Maksimovic, J., Gordon, L., & Oshlack, A. (2012). SWAN: Subset-quantile within array normalization for illumina infinium HumanMethylation450 BeadChips. Genome biology, 13(6), 1–12.

Marioni, R. E., Shah, S., McRae, A. F., Chen, B. H., Colicino, E., Harris, S. E., . . . Cox, S. R. (2015). DNA methylation age of blood predicts all-cause mortality in later life. Genome biology, 16(1), 25.

Marioni, R. E., Shah, S., McRae, A. F., Ritchie, S. J., Muniz-Terrera, G., Harris, S. E., . . . Pattie, A. (2015). The epigenetic clock is correlated with physical and cognitive fitness in the Lothian Birth Cohort 1936. International journal of epidemiology, 44(4), 1388–1396.

Mendelson, M. M., Marioni, R. E., Joehanes, R., Liu, C., Hedman, Å. K., Aslibekyan, S., . . . Yao, C. (2017). Association of body mass index with DNA methylation and gene expression in blood cells and relations to cardiometabolic disease: a Mendelian randomization approach. PLoS medicine, 14(1).

Michailidou, K., Lindström, S., Dennis, J., Beesley, J., Hui, S., Kar, S., . . . Rostamianfar, A. (2017). Association analysis identifies 65 new breast cancer risk loci. Nature, 551(7678), 92.

Nikpay, M., Goel, A., Won, H.-H., Hall, L. M., Willenborg, C., Kanoni, S., . . . Hopewell, J. C. (2015). A comprehensive 1000 Genomes–based genome-wide association meta-analysis of coronary artery disease. Nature genetics, 47(10), 1121.

Palmer, T. M., Lawlor, D. A., Harbord, R. M., Sheehan, N. A., Tobias, J. H., Timpson, N. J., . . . Sterne, J. A. (2012). Using multiple genetic variants as instrumental variables for modifiable risk factors. Statistical methods in medical research, 21(3), 223–242.

Patterson, D. G., Roberts, J. T., King, V. M., Houserova, D., Barnhill, E. C., Crucello, A., . . . Nguyen, M. (2017). Human snoRNA-93 is processed into a microRNA-like RNA that promotes breast cancer cell invasion. NPJ Breast Cancer, 3(1), 1–12.

Phelan, C. M., Kuchenbaecker, K. B., Tyrer, J. P., Kar, S. P., Lawrenson, K., Winham, S. J., . . . Chornokur, G. (2017). Identification of 12 new susceptibility loci for different histotypes of epithelial ovarian cancer. Nature genetics, 49(5), 680.

Pidsley, R., Wong, C. C., Volta, M., Lunnon, K., Mill, J., & Schalkwyk, L. C. (2013). A data-driven approach to preprocessing Illumina 450K methylation array data. BMC genomics, 14(1), 1–10.

Pilling, L. C., Kuo, C.-L., Sicinski, K., Tamosauskaite, J., Kuchel, G. A., Harries, L. W., . . . Melzer, D. (2017). Human longevity: 25 genetic loci associated in 389,166 UK biobank participants. Aging (Albany NY), 9(12), 2504.

Richard, M. A., Huan, T., Ligthart, S., Gondalia, R., Jhun, M. A., Brody, J. A., . . . Tsai, P.-C. (2017). DNA methylation analysis identifies loci for blood pressure regulation. The American Journal of Human Genetics, 101(6), 888–902.

Schumacher, F. R., Al Olama, A. A., Berndt, S. I., Benlloch, S., Ahmed, M., Saunders, E. J., . . . Cieza-Borrella, C. (2018). Association analyses of more than 140,000 men identify 63 new prostate cancer susceptibility loci. Nature genetics, 50(7), 928.

Scott, R. A., Scott, L. J., Mägi, R., Marullo, L., Gaulton, K. J., Kaakinen, M., . . . Eicher, J. D. (2017). An expanded genome-wide association study of type 2 diabetes in Europeans. Diabetes, 66(11), 2888–2902.

Shah, R. V., Hwang, S.-J., Yeri, A., Tanriverdi, K., Pico, A. R., Yao, C., . . . Demarco, D. (2019). Proteins altered by surgical weight loss highlight biomarkers of insulin resistance in the community. Arteriosclerosis, thrombosis, and vascular biology, 39(1), 107–115.

Svane, A. M., Soerensen, M., Lund, J., Tan, Q., Jylhävä, J., Wang, Y., . . . Deary, I. J. (2018). DNA methylation and all-cause mortality in middle-aged and elderly Danish twins. Genes, 9(2), 78.

Teschendorff, A. E., Marabita, F., Lechner, M., Bartlett, T., Tegner, J., Gomez-Cabrero, D., & Beck, S. (2013). A beta-mixture quantile normalization method for correcting probe design bias in Illumina Infinium 450 k DNA methylation data. Bioinformatics, 29(2), 189–196.

Timmers, P. R., Mounier, N., Lall, K., Fischer, K., Ning, Z., Feng, X., . . . Esko, T. (2019). Genomics of 1 million parent lifespans implicates novel pathways and common diseases and distinguishes survival chances. eLife, 8, e39856.

Triche Jr, T. J., Weisenberger, D. J., Van Den Berg, D., Laird, P. W., & Siegmund, K. D. (2013). Low-level processing of Illumina Infinium DNA methylation beadarrays. Nucleic acids research, 41(7), e90–e90.

van den BeCrg, N., Beekman, M., Smith, K. R., Janssens, A., & Slagboom, P. E. (2017). Historical demography and longevity genetics: back to the future. Ageing research reviews, 38, 28–39.

Van Eyken, E., Van Laer, L., Fransen, E., Topsakal, V., Lemkens, N., Laureys, W., . . . Van De Heyning, P. (2006). KCNQ4: a gene for age-related hearing impairment? Human mutation, 27(10), 1007–1016.

Walter, S., Atzmon, G., Demerath, E. W., Garcia, M. E., Kaplan, R. C., Kumari, M., . . . Tranah, G. J.(2011). A genome-wide association study of aging. Neurobiology of aging, 32(11), 2109. e2115–2109. e2128.

Wang, Y., McKay, J. D., Rafnar, T., Wang, Z., Timofeeva, M. N., Broderick, P., . . . Han, Y. (2014). Rare variants of large effect in BRCA2 and CHEK2 affect risk of lung cancer. Nature genetics, 46(7), 736.

Willer, C. J., Schmidt, E. M., Sengupta, S., Peloso, G. M., Gustafsson, S., Kanoni, S., . . . Mora, S. (2013). Discovery and refinement of loci associated with lipid levels. Nature genetics, 45(11), 1274.

Yao, C., Chen, G., Song, C., Keefe, J., Mendelson, M., Huan, T., . . . Wu, H. (2018). Genome-wide mapping of plasma protein QTLs identifies putatively causal genes and pathways for cardiovascular disease. Nature Communications, 9(1), 1–11.

Zhang, Y., Wilson, R., Heiss, J., Breitling, L. P., Saum, K.-U., Schöttker, B., . . . Brenner, H. (2017). DNA methylation signatures in peripheral blood strongly predict all-cause mortality. Nature Communications, 8(1), 1–11.

Zhao, Y., Yan, Y., Ma, R., Lv, X., Zhang, L., Wang, J., . . . Zhao, L. (2020). Expression signature of six-snoRNA serves as novel non-invasive biomarker for diagnosis and prognosis prediction of renal clear cell carcinoma. Journal of cellular and molecular medicine, 24(3), 2215–2228.

