## Additional File 1 for "An integrative analysis of clinical and epigenetic biomarkers of mortality"

**Additional file1**

**Cohorts Description**

**Atherosclerosis Risk in Communities Study (ARIC)**

**Study participants:** The Atherosclerosis Risk in Communities (ARIC) study is an ongoing multicenter prospective cohort study that enrolled participants in four communities: Forsyth County, NC; Jackson, MS; northwest suburbs of Minneapolis, MN; and Washington County, MD. ARIC is designed to investigate the etiologies of atherosclerosis and its related diseases, as well as variation in cardiovascular disease risk factors, medical care, and disease by race, sex, location, and date(Investigators, 1989b). ARIC consists of 15,792 individuals aged 45-64. Clinic visits were conducted in 1987-89, 1990-92, 1993-95, 1996-98, 2011-13, 2016-17, and 2018-19. The Institutional Review Boards at study centers approved study protocols.

**Mortality ascertainment and clinical phenotypes:** Deaths prior to January 1, 2017 were ascertained through annual telephone interviews with participants or their next of kin, surveillance of local hospital discharge records and state death records, and linkage to the National Death Index(Investigators, 1989a; White et al., 1996). Date and cause of death was ascertained from death certificate review.

Trained study staff measured anthropometrics including height and weight using standard protocols with participants lightly dressed and not wearing footwear(Investigators, 1989a). BMI was calculated as weight in kilograms divided by the square of height in meters. Questionnaires ascertained participant education in years, smoking history, and alcohol use(Investigators, 1989a). The Baecke questionnaire measured participant physical activity(Investigators, 1989a; Richardson, Ainsworth, Wu, Jacobs Jr, & Leon, 1995). Prevalent CVD status was defined by self-report of physician-diagnosed myocardial infarction (MI), evidence of silent MI by ECG, history of coronary revascularization, or self-report of physican-diagnosed stroke(White et al., 1996). Prevalent type 2 diabetes status was defined as participant reporting medications use for type 2 diabetes, self report of type 2 diabetes diagnosed by a physician, fasting blood glucose of at least 126 mg/dL, or non-fasting blood glucose of at least 200 mg/dL. Prevalent hypertension status was defined as participant report of antihypertensive medication use, systolic blood pressure of at least 140 mmHg, or diastolic blood pressure of at least 90 mmHg. Cancer was ascertained through linkage with cancer registries and surveillance of local hospital records, described previously(Rasmussen-Torvik et al., 2013). Analyses excluded cases of non-melanoma skin cancers.

**DNA methylation measurements and quality control:** DNA from peripheral blood leukocytes were extracted with the Gentra Puregene Blood Kit (Qiagen; Valencia, CA, USA) per manufacturer instructions ([www.qiagen.com](http://www.qiagen.com)). DNA was obtained from samples collected at either visit 2 (1990-92) or visit 3 (1993-95), which serve as the baseline for mortality follow-up. The deep well format EZ-96 DNA Methylation Kit (Zymo Research; Irvine, CA, USA) bisulfite-converted the extracted DNA per manufacturer instructions ([www.zymoresearch.com](http://www.zymoresearch.com)). The bisulfite-converted DNA then underwent whole genome amplification, fragmentation, hybridization, and single base-pair extension with a fluorescently labeled nucleotide and subsequently scanned. The Illumina Infinium Human Methylation 450 BeadChip (HM450K) measured DNA methylation. DNA methylation in African American and European American participants were measured in separate batches. Image intensities were extracted using the Illumina GenomeStudio software and the DNAm β values were processed using the normal exponential convolution using out of band probes (noob) technique(Triche Jr, Weisenberger, Van Den Berg, Laird, & Siegmund, 2013). The noob-processed DNA methylation data was the normalized using beta mixture quantile dilation (BMIQ)(Teschendorff, Marabita, et al., 2012).

**Epigenome-wide association analysis of mortality:** We used Cox proportional hazards regression for time-to-event analyses to test associations of CpG beta values with all-cause mortality, CVD mortality, and cancer mortality using the coxph function in the R “survival” package. Models adjusted for clinical phenotypes, covariates, white blood cell count, white blood cell type proportions, and Illumina HM450K control probe principal components to account for batch effects(Benjamin Lehne, Alexander W Drong, et al., 2015).

**The Cardiovascular Health Study (CHS)**

**Study participants:** The CHS is a population-based cohort study of risk factors for coronary heart disease and stroke in adults ≥65 years conducted across four field centers(Fried et al., 1991). The original predominantly European ancestry cohort of 5,201 persons was recruited in 1989-1990 from random samples of the Medicare eligibility lists; subsequently, an additional predominantly African-American cohort of 687 persons was enrolled for a total sample of 5,888. CHS was approved by institutional review committees at each field center and individuals in the present analysis had available DNA and gave informed consent including consent to use of genetic information for the study of cardiovascular disease.

**DNA methylation measurements and quality control:** DNA methylation was measured on a randomly selected subset of 336 European ancestry and 329 African-American ancestry participants who participated in the 3rd annual follow-up visit (study year 5) and had DNA available from that visit.  The European ancestry participants had no baseline history of coronary vascular disease (defined as coronary heart disease, congestive heart failure, peripheral vascular disease, valvular heart disease, stroke, or transient ischemic attack).

Methylation measurements were performed at the Institute for Translational Genomics and Population Sciences at the Harbor-UCLA Medical Center Institute for Translational Genomics and Population Sciences (Los Angeles, CA).   DNA was extracted from Buffy coat fractions and subsequently underwent bisulfite conversion using the EZ DNA Methylation kit (Zymo Research, Irvine, CA).  Methylation was then assayed using the Infinium HumanMethylation450 BeadChip (Illumina Inc, San Diego, CA).

Quality control was performed in in the minfi R package (version 1.12.0, <http://www.bioconductor.org/packages/release/bioc/html/minfi.html)>(Martin J Aryee et al., 2014; Fortin et al., 2014). Samples with low median intensities of below 10.5 (log2) across the methylated and unmethylated channels, samples with a proportion of probes falling detection of greater than 0.5%, samples with QC probes falling greater than 3 standard deviation from the mean, sex-check mismatches, failed concordance with prior genotyping or > 0.5% of probes with a detection p-value > 0.01 were removed. Probes with >1% of values below detection were removed.  In total, 11 samples were removed for sample QC resulting in a sample of 323 European-ancestry and 326 African-American samples.   Methylation values were normalized using the SWAN quantile normalization method(Maksimovic, Gordon, & Oshlack, 2012).  Since white blood cell proportions were not directly measured in CHS they were estimated from the methylation data using the Houseman method(Eugene Andres Houseman et al., 2012).

**Mortality ascertainment and clinical phenotypes:** Deaths among CHS participants that occurred during the 10 years following the 1992-93 study visit were identified during surveillance calls, during scheduling calls for annual clinic visits, using data from the Centers for Medicare & Medicaid Services and the National Death Index, and through review of newspaper obituaries. Cause of death was adjudicated by a committee of physicians using standardized criteria to review all available evidence including death certificates, autopsy/coroner reports, hospital medical records, and interview with the decedent’s next-of-kin or caregiver.

Height, and weight were measured using standard protocols. BMI was calculated as weight (kg) per height squared (m2). Age, years of education, smoking, alcohol drinking and history of cancer were based on self-report and assessed by interviewer-administered questionnaire. Physical activity (kcal/wk) was assessed using a modified Minnesota Leisure-Time Activities questionnaire(Taylor et al., 1978). Diabetes was defined as measured fasting blood glucose >125 mg/dL or use of diabetes medication. Hypertension was defined as systolic blood pressure (BP) ≥ 140 mmHg, diastolic BP ≥90 mmHg, or use of hypertension medication plus a physician diagnosis of hypertension.

**The Danish Twin Registry sample (DTR)**

**Study participants:** The study population was comprised of 870 twin individuals (435 pairs of monozygotic twins) from three different population-based nation-wide twin surveys conducted within the frames of the Danish Twin Registry(Pedersen et al., 2019): the study of birth weight-discordant twins (BWD), the Study of Middle-Aged Danish Twins (MADT) and the Longitudinal Study of Aging Danish Twins (LSADT). In the present study 150 individuals from BWD, 482 individuals from MADT and 238 individuals from LSADT were included, similarly to Svane et al. 2018(Svane et al., 2018). Blood samples were drawn over the periods 2008-2011 for MADT and BWD, and 1996–1997 for LSADT. Informed consents were obtained from all participants and all surveys were approved by The Regional Committees on Health Research Ethics for Southern Denmark (S-VF-19980072, S-VF-20040241 and S-20090033) and were conducted in accordance with the Helsinki II declaration.

**Mortality ascertainment and clinical phenotypes:** Dates of birth and dates of death were obtained from the Danish Civil Registration System(Mortensen, Gøtzsche, Bøcker Pedersen, & Østrup Møller, 2006) per 1^st^ of March 2019, while causes of death were obtained from the National Causes of Death Registry per 31^st^ of December 2012. All three twin surveys included comprehensive face-to-face interviews and assessments of numerous health outcomes(Pedersen et al., 2019). For the present study self-reported ever occurrence of diseases (cancer, diabetes, treated hypertension, stroke, angina pectoris, heart attack, irregular heart rhythm and general heart problems), and life style habits (smoking (never/former vs. current smoker), alcohol intake (g/day), and level of physical activity (frequency and intensity)), as well as level of education (years in school) were included. Finally, bmi was calculated (kg/m^2^) based on either measured weight and height (BWD and MADT) or measured or estimated weight and height (LSADT).

**DNA methylation measurement and quality control:** DNA was isolated from buffy coat by salt precipitation, using either a manual protocol or a semi-automated protocol with the Autopure System (Qiagen, Hilden, Germany). Bisulfite conversion of 500 ng genomic DNA was conducted with the EZ Methylation Gold Kit (Zymo Research, Orange County, CA, USA). The methylation level of 485,512 CpG sites was analysed by the Infinium 450K HumanMethylation BeadChip (Illumina, San Diego, CA, USA) following the manufacturer’s instructions at either the Leiden University Medical Center or GenomeScan B.V., Leiden, the Netherlands. BeadChip images were scanned using the iScan system. Twin pairs were analysed on the same array. The BWD, MADT and LSADT samples were analysed on different occasions. Data pre-processing and quality control were performed using the free R packages MethylAid(Van Iterson et al., 2014) and minfi(Martin J Aryee et al., 2014; Fortin et al., 2014) for each dataset as described in Soerensen et al. 2019(Soerensen et al., 2019). In short, samples were excluded if less than 95% of the probes had a detection P-value<0.01, or if the samples failed based on the internal quality control probes used by MethylAid(Van Iterson et al., 2014)^.^ Probes were excluded if they had a detection P-value>0.01, a raw intensity value of zero, a low bead count (<3 beads), a measurement success rate below 95% or had been identified as being cross-reactive^7.^ The DNA methylation data were annotated using GRCh37/hg19 using the annotation file and recommendations supplied by Illumina Inc. (Illumina, San Diego, CA, United States). In total 451,471 CpGs were available for analysis after quality control for the present study. Normalization was carried out by Functional normalization^8^.

**Epigenome-wide association analysis of mortality:** In order to eliminate unwanted variation in the DNA methylation data, a principle component analysis (PCA) was conducted before epigenome-wide association analysis (EWAS), similarly to Tobi et al. 2015(Tobi et al., 2015); technical variables reflecting bathes, plate number, position on array etc., as well as cell counts were included. Blood cell type composition had previously been measured using a Coulter LH 750 Haematology Analyser for most MADT and BWD individuals and these values were used for imputing the proportion of basophils, eosinophils, monocytes, neutrophils, and lymphocytes in the remaining individuals (see Debrabant et al. 2017(Debrabant et al., 2018) for details). The PCA revealed PCs 1-4 to describe the majority of the variance in the data (the subsequent PCs accounted for less than 3% of the variance), consequently PCs 2-4 were included in the EWAS. PC1 was not included as it reflected much of the same variance as sex. Furthermore, in order to avoid over adjustment, cell counts and technical variables were not included in the EWAS as they either did not reflect the variance in the data or reflected much of the same variance as PC3.

Epigenome-wide association analysis was performed using rank-based inversed normal transformed DNA methylation via Cox proportional hazards models to test for associations between each CpG and mortality outcomes including all-cause death, CVD death and cancer death. For all-cause mortality, survival until the 1st of March 2019 was investigated, while for cause-specific mortality, survival until the 31st of December 2012 was investigated. The coxph() function in the ‘survival’ R library was applied. The analysis was adjusted for age, sex, PCs2-4, and familiar relatedness (twin pair structure) in one model, and additionally adjusting for the clinical and lifestyle phenotypes mentioned above (cf. the section on Mortality ascertainment and clinical phenotypes).

**The ESTHER Study**

**Study participants:** The ESTHER study is an ongoing population-based cohort study conducted in the federal state of Saarland, Germany.(Raum et al., 2007) In brief, 9,949 older adults (50-75 years) were recruited by their general practitioners (GPs) during routine health check-ups (offered every two years to people older than 35 years in the German healthcare system) between 2000 and 2002, and followed up thereafter. A subset of 1000 ESTHER participants who were consecutively enrolled during the first 3 months of recruitment was selected for DNA methylation assessment in the baseline blood samples and was included in the current study.(Zhang et al., 2017) The study was approved by the ethics committees of the University of Heidelberg and of the Medical Association of Saarland. All participants provided written informed consent.

**Mortality ascertainment and clinical phenotypes:** Deaths of ESTHER participants during follow-up were identified through record linkage with population registries in Saarland. Information on the major cause of death was obtained from death certificates provided by the local health authorities, and coded with ICD-10-codes. Deaths from CVD and malignant invasive cancers, respectively, were defined by ICD-10 codes I00-I99 and C00-C97 (excluding non-melanoma skin cancer (C44)).

During the baseline enrolment, epidemiological data (including socio-demographic characteristics, lifestyle factors, and history of major diseases) were collected via a standardized self-administered questionnaire completed by participants and via additional reports from participants’ GPs, and biological samples (blood, stool, urine) were obtained and stored at −80 °C.(Zhang et al., 2017)

**DNA methylation measurements and quality control:** DNA from the whole blood collected at baseline was extracted using a salting out procedure,(Miller, Dykes, & Polesky, 1988) and allocated in the 96-well format, where 3 random duplicate samples were placed as quality controls. DNAm in whole blood was quantified using the Infinium HumanMethylation450K BeadChip (Illumina.Inc, San Diego, CA, USA) at the Genomics and Proteomics Core Facility of the German Cancer Research Center, Heidelberg, Germany. In brief, 1.5mg DNA was bisulfite converted (Zymo Research, Irvine, CA), and 200ng bisulfite-treated DNA was applied to the 450K BeadChips following the manufacturer’s instruction. Raw data pre-processing and initial quality control was carried out following the CPACOR pipeline.(Benjamin Lehne, Alexander W Drong, et al., 2015) Probes with detection p-value>0.01 and targeting the sex chromosomes, cross-reactive probes, and polymorphic CpGs(Chen et al., 2013) were removed before quantile normalization, which was applied following stratification of the probe type into categories according to probe type and colour channel, using the R package limma. Sample call rate threshold and CpG call rate threshold both were 95%. A total of 995 samples and 417,887 CpGs were retained for EWAS analysis. A principle component analysis (PCA) was performed for the positive control probes, and the first 20 control probe PCAs were included in the regression model as technical covariates.

**Epigenome-wide association analysis of mortality:** Cox proportional hazards regression models were fitted to estimate the associations of methylation beta values with all-cause, CVD, and cancer mortality, using *coxph()* function in the ‘survival’ R package. Methylation beta values were transformed by rank-based inverse normal transformation before entering the regression models.

**The Framingham Heart Study (FHS)**

**Study participants:** The FHS is a community-based study^(Dawber, Meadors, & Moore Jr, 1951)^. In 1971, the offspring and their spouses of the FHS original cohort were recruited(Feinleib, Kannel, Garrison, McNamara, & Castelli, 1975). In this study, eligible participants included offspring cohort attendees at their eighth examination cycle (Exam 8, 2005-2008, N=2427). The study protocol was approved by the Institutional Review Board at Boston University Medical Center (Boston, MA).

**Mortality ascertainment and clinical phenotypes:** Deaths among FHS participants that occurred prior to January 1, 2016 were ascertained using multiple strategies, including routine contact with participants for health history updates, surveillance at the local hospital and in obituaries of the local newspaper, and queries to the National Death Index. Death certificates, hospital and nursing home records prior to death, and autopsy reports were requested. When cause of death was undeterminable, the next of kin were interviewed. The date and cause of death were reviewed by an endpoint panel of 3 investigators.

Age, height, and weight were measured using standard protocols. BMI was calculated as weight (kg) per height squared (m^2^). Education years, physical activity, smoking, alcohol drinking were self-reported traits ascertained by physician-administered questionnaire. An individual was defined as having CVD if he or she had coronary heart disease or stroke. Diabetes was defined as either use of diabetes medication or a measured fasting blood glucose level of >125 mg/dL. Hypertension was defined as either use of antihypertensive treatment or a measure of systolic blood pressure (BP) ≥ 140 mmHg or diastolic BP ≥90 mmHg. A cancer phenotype was defined if the person had any type of cancer excluding non-melanoma skin cancer.

**DNA methylation measurements and quality control:** DNA samples were extracted from whole blood buffy coat samples using the Gentra Puregene DNA extraction kit (Qiagen, Venlo, Netherland) and subsequently underwent bisulfite conversion using EZ DNA methylation kit (Zymo Research, Irvine, CA). Samples underwent whole genome amplification, fragmentation, array hybridization, and single-base pair extension. DNA methylation levels were measured using Illumina Infinium Human Methylation450 BeadChip (450K). FHS offspring cohort samples were run in two laboratory batches at the Johns Hopkins Center for Inherited Disease Research (lab batch #1) and University of Minnesota Biomedical Genomics Center (lab batch #2). For each separately lab batch, DNAm β values from Illumina GenomeStudio were further normalized using the DASEN methodology implemented in the wateRmelon R package and output the final β values of each CpG for downstream analysis(Pidsley et al., 2013). DNAm β values indicate the proportion of DNA molecular in the sample that were methylated at the same CpG site, ranging from 0 to 1. For sample quality control, we excluded samples with a methylation value missing (detection P >0.01) at >1% CpGs, poor matching of single nucleotide polymorphisms (SNPs) between the 65 SNPs on the Illumina 450K array and the GWAS array, or outliers at the multi-dimensional scaling plot. For quality control at the probe level, we excluded probes with methylation value missing (detection P>0.01) at >20% samples, as well as probes previously identified to map to multiple locations(Price et al., 2013) on the sex chromosomes, or to have an underlying SNP (minor allele frequency [MAF] >5% in 1000 Genomes Project data) at the CpG site or within 10bp of the single base extension(Chen et al., 2013). A total of 415,318 CpGs were retained for further analysis.

**Epigenome-wide association analysis of mortality:** We used surrogate variable analyses (SVA) to eliminate unwanted variation in the DNAm data(Leek & Storey, 2007). SVs were generated in each lab batch separately. DNAm beta values were regressed on batch-specific SVs, and the DNAm residual was taken forward. The two lab batches were merged together for analysis. In our previous studies using the same data, the SVA method was demonstrated to adequately account for latent batch effects, variations in blood cell proportions, and technical covariates(Liu et al., 2016; Mendelson et al., 2017). Therefore, to avoid over adjustment, we did not further adjust for technical covariates (i.e. plates, rows and columns) or cell types in DNAm data. We performed rank-based inverse normal transformation of DNAm residual (after adjusted for SVs). Then, we conducted time-to-event analyses using Cox proportional hazards models to test for associations between each CpG and mortality outcomes including all-cause death, CVD death and cancer death, using *coxph*() function in the ‘survival’ R library, adjusting for clinical phenotypes and familiar relatedness.

**Invecchiare in Chianti, aging in the Chianti area (InCHIANTI study)**

**Study participants:** The Invecchiare in Chianti (InCHIANTI) Study is a population-based prospective cohort study of residents ages 20 or older from two areas in the Chianti region of Tuscany, Italy. Sampling and data collection procedures have been described elsewhere.(Ferrucci et al., 2000) Briefly, blood sample for DNA extraction was collected at baseline (1998-2000). All participants provided written informed consent to participate in this study. The study complied with the Declaration of Helsinki. The Italian National Institute of Research and Care on Aging Institutional Review Board approved the study protocol.

**Mortality ascertainment and clinical phenotypes:** Data on demographic and lifestyle factors such as smoking, years of education and physical activity were collected during the baseline interview. Physical activity in the previous year was assessed as sedentary (hardly any physical activity, ii) mostly sitting/some walking, iii) Light exercise 2-4hrs/week, iv) moderate 1-2 hours/week, v) moderate exercise > 3hrs/week, vi) intensive exercise many times/wk and vii) walk over 5km/day for 5 days for at least 5 years. For the analysis, the categories were collapsed into sedentary (i-ii) versus non-sendentary (iii-vi). Smoking was categorized into current smoker versus former and non-smokers. Weight and height were measured by using standard techniques. Body mass index (BMI) was calculated as measured weight in kilograms divided by measured height in meters squared (kg/m2). Alcohol consumption (in gram/day) during the past year was assessed using a 236 item food frequency questionnaire (FFQ) for the European Prospective Investigation on Cancer and nutrition (EPIC) study, previously validated in the InCHIANTI population (Pisani et al., 1997). Chronic disease status at baseline including hypertension, cardiovascular disease, stroke, heart failure, type 2 diabetes, and cancer were defined using standard criteria that combined information from self-reported medical history, medication use, medical documents, and a clinical medical examination. Vital status was ascertained using data from the Tuscany Regional Mortality General Registry. Deaths were assessed until

**DNA methylation measurements and quality control:** Genomic DNA was extracted from buffy coat samples using an AutoGen Flex and quantified on a Nanodrop1000 spectrophotometer prior to bisulfite conversion. Genomic DNA was bisulfite converted using Zymo EZ-96 DNA Methylation Kit (Zymo Research Corp., Irvine, CA) as per the manufacturer’s protocol. CpG methylation status of 485,577 CpG sites was determined using the Illumina Infinium HumanMethylation450 BeadChip (Illumina Inc., San Diego, CA) as per the manufacturer’s protocol and as previously described.(Moore et al., 2016) Initial data analysis was performed using GenomeStudio 2011.1 (Model M Version 1.9.0, Illumina Inc.). Threshold call rate for inclusion of samples was 95%. Quality control of sample handling included comparison of clinically reported sex versus sex of the same samples determined by analysis of methylation levels of CpG sites on the X chromosome.(Moore et al., 2016) Background subtraction was applied using the preprocessIllumina command in the minfi Bioconductor package(M. J. Aryee et al., 2014).

**Epigenome-wide association analysis of mortality:** Mortality analysis was conducted using cox proportional hazards regression models to estimate the associations of methylation beta values with all-cause, CVD, and cancer mortality, using coxph() function in the ‘survival’ R package. Each methylation beta values were inverse rank normalized. The analyses were adjusted for age at baseline, sex, alcohol consumption, smoking, BMI, physical activity, education, chronic diseases (hypertension, type 2 diabates, cancer, stroke, heart failure) and assay batch.

**The KORA study (Cooperative health research in the Region of Augsburg)**

**Study participants:** KORA is an independent population-based cohort sampled from the region of Augsburg, Southern Germany(Holle, Happich, Löwel, & Wichmann, 2005). The study has been conducted according to the principles of the Declaration of Helsinki. All participants provided written informed consent. The local ethics committee (Bayerische Landesärztekammer) reviewed and approved the study. All participants underwent detailed physical examination with blood sample collection and completed questionnaires to gather clinical and socio-demographic information. For this study, we used data of the KORA-F4 survey participants with methylation data which was conducted (2006-2008, N = 1727).

**Mortality ascertainment and clinical phenotypes:** Mortality among the participants of the KORA F4 study was ascertained using death certificates, coded according to the International Classification of Diseases (originally ICD-9, translated to ICD-10) (Schederecker et al., 2020). Cardiovascular disease (CVD)-related mortality was identified using the codes (ICD-9 codes 390 – 459, ICD-10 codes I00–I99) for diseases of the cardiovascular system and the code (ICD-9 code 798, ICD-10 code R99) for sudden death with unknown cause. Cancer-related mortality was identified using the codes (ICD-9 codes 140–208, ICD-10 codes C00–C95). All-cause mortality included CVD-related mortality, cancer-related mortality as well as other disease-related mortality like pneumonia (ICD-9 code 486, ICD-10 codes J18.8, J18.9), chronic bronchitis (ICD-9 code 491, ICD-10 Codes J41, J42, J44) and dementias (ICD-9 code 290, ICD-10 Codes F03.90, F05, F01.50, F01.51). Age, height, and weight were measured during physical examination with BMI calculated as weight (kg) per height squared (m2). Education years, physical activity, smoking status, alcohol consumption were self-reported. Coronary heart disease, heart failure, stroke and cancer were self-reported. Diabetes was defined as self-reported disease status or the use of anti-diabetic medication. Hypertension was defined having a measure of systolic blood pressure (BP) ≥ 140 mmHg or diastolic BP ≥90 mmHg or having medically controlled hypertension.

**DNA methylation measurements and quality control:** Genome-wide DNA methylation measurement at 485,577 genomic sites was performed using the Infinium HumanMethylation450K BeadChip® (Illumina, Inc., CA, USA)(Bibikova et al., 2011) in 1802 KORA F4 samples with laboratory process as described previously(Zeilinger et al., 2013).

DNA methylation data were preprocessed following the CPACOR pipeline of Lehne et al.(Benjamin Lehne, Alexander W. Drong, et al., 2015). First, 65 probes that represent SNPs were excluded. Second, background correction was performed using the R package minfi, version 1.6.0(M. J. Aryee et al., 2014). Third, detection p-values were defined as the probability of a signal being detected above the background signal level, as estimated from negative control probes. Consequently, signals with detection p-values ≥ 0.01 and signals summarized from less than three functional beads on the chip were removed, as they are putatively unreliable. To reduce the non-biological variability between observations, data were normalized using quantile normalization on the raw signal intensities. Precisely, QN was performed on a stratification of the probe categories into 6 types, based on probe type and color channel, using the R package limma, version 3.16.5(Smyth, 2005). Following normalization, for each CpG site methylated and unmethylated signal intensities were converted to β-values, the ratio of the methylated signal intensity divided by the overall signal intensity(Bibikova et al., 2011; Du et al., 2010): β-value = M/(M+U+α), where an offset was added as a regularization for the situation when both M and U are low, as recommended by Illumina(Du et al., 2010).To further reduce technical variation, the principal components of the non-negative methylation control probes were calculated. The first 30 principal components were used as covariates in the regression models, as recommended by Lehne et al(Benjamin Lehne, Alexander W. Drong, et al., 2015).

White blood cell proportions are important determinants of methylation in whole blood. Since cell type measurements were not available for the KORA data, proportions of selected cell types (i.e., granulocytes, monocytes, B cells, CD4+ T cells, CD8+ T cells and natural killer cells) were estimated using the procedure proposed by Houseman et al.(Eugene Andres Houseman et al., 2012), and included as covariates in the models.

**Epigenome-wide association analysis of mortality:** We performed time-to-event analyses using Cox proportional hazards models to test for associations between each inverse normal transformed CpG and mortality outcomes including all-cause death, CVD death and cancer death, using *coxph*() function in the ‘survival’ R library, adjusting for clinical phenotypes, the first 30 principal components of methylation data and Houseman estimated cell types.

**Normative Aging Study (NAS)**

**NAS cohort description.** The ongoing longitudinal US Department of Veterans Affairs NAS was established in 1963 and included men 21–80 years old and free of known chronic medical conditions at entry.(Bell, Rose, & Damon, 1966) Participants were invited to medical examinations every three to five years. At each visit, men provided information on medical history, lifestyle, and demographic factors and underwent physical examinations and laboratory tests. DNA samples were collected from 675 active participants between 1999–2007.(Bell et al., 1966) We excluded participants who were non-white or who reported leukemia at the time of DNA extraction, leaving a total of 646 individuals with a single observation each. Participants provided written informed consent at each visit. The NAS study was approved by the institutional review boards of participating institutions. At each in-person visit, participants completed questionnaires regarding demography, lifestyle, and medical history. They reported chronological age, years of education, smoking status (never, former, current), pack-years consumption (continuous), alcohol consumption (<2, ≥2 drinks/day), physical activity (<12, 12–30, ≥30 metabolic equivalent hours [MET-h] per week), type 2 diabetes (self-reported diagnosis and/or use of diabetes medications), diagnosis of CHD (validated on medical records, ECG, and physician exams), diagnosis of malignant cancer in the five years prior the visit (diagnosed with ICD-9 code). High blood pressure was defined as antihypertensive medication use, systolic blood pressure ≥140 mmHg, or diastolic blood pressure ≥90 mmHg at study visit. BMI was computed from anthropometric measures, performed with participants in undershorts and socks.(Troisi, Heinold, Vokonas, & Weiss, 1991)

**NAS mortality ascertainment.** Official death certificates were obtained for decedents from the appropriate state health departments and were reviewed by a physician. An experienced research nurse coded the cause of death using ICD-9. Both participant deaths and causes of death were routinely updated by the research team, and the last update available was December 31, 2013.(Marioni et al., 2015)

**DNA methylation measures.** DNA was extracted from buffy coats using the QIAamp DNA Blood Kit (Qiagen). We used 500 ng of DNA for bisulfite conversion using the EZ-96 DNA Methylation Kit (Zymo Research). To reduce chip and plate effects, we used a two-stage age-stratified algorithm to randomize samples and ensure similar age distributions across chips and plates; 12 samples that were sampled across all age quartiles were randomized to each chip, and then chips were randomized to plates (8 chips/plate). QC analysis was performed to remove samples and probes, where >1% of probes or samples, respectively, had a detection P > 0.05. Remaining samples were preprocessed using the Illumina-type background correction(Triche Jr et al., 2013) and normalized with dye-bias(Davis, Du, Bilke, Triche, & Bootwalla, 2017) and BMIQ(Teschendorff, Jones, et al., 2012) adjustments, which were used to generate beta methylation values. The working set included 477, 928 CpG probes. DNA methylation age was computed using the Horvath calculator from background-corrected methylation data, and QC analysis was performed only on samples, leaving 485, 512 CpG and CpH probes in the working set.

**Rotterdam Study (RS)**

**Study population:** The Rotterdam Study (RS) is a large prospective, population-based cohort study aimed at assessing the occurrence of and risk factors for chronic (cardiovascular, endocrine, hepatic, neurological, ophthalmic, psychiatric, dermatological, oncological, and respiratory) diseases in the elderly (Ikram et al., 2020). The study comprises 14,926 subjects in total, living in the well-deﬁned Ommoord district in the city of Rotterdam in the Netherlands. In 1989, the first cohort, Rotterdam Study-I (RS-I) comprised of 7,983 subjects with age 55 years or above. In 2000, the second cohort, Rotterdam Study-II (RS-II) was included with 3,011 subjects who had reached an age of 55 or over in 2000. In 2006, the third cohort, Rotterdam Study-III (RS-III) was further included with 3,932 subjects with age 45 years and above. Each participant gave an informed consent and the study was approved by the medical ethics committee of the Erasmus University Medical Center, Rotterdam, the Netherlands.

**Mortality ascertainment and other phenotypes:** Data collection procedures have been described elsewhere(Ikram et al., 2020). Briefly, information on vital status was collected weekly from municipal population registries as well as from general practitioners and hospitals records. All-cause mortality was defined as participants who died from any cause until January 2017.

Data on highest education level, smoking status, physical activity and alcohol consumption were obtained through self-report during home interviews. Information on physical activity was collected using questionnaires Zutphen(Caspersen, Bloemberg, Saris, Merritt, & Kromhout, 1991) (RS-I and RS-II) and LASA(Stel et al., 2004) (RS-III), and expressed in MET-hours/week. Physical measures and collection of blood samples were assessed during visits to the research centers, using standard procedures(Ikram et al., 2020). BMI was calculated as weight (kg) divided by height (meters) squared. Disease information was collected by self-report and by general practitioner and pharmacy records. Disease events were coded according to the International Classification of Diseases 10th version (ICD-10) by two independent research physicians. Cardiovascular disease cases were defined as the occurrence of coronary heart disease or stroke. Type 2 diabetes was defined according to the current WHO guidelines: fasting blood glucose ≥7.0 mmol/L, non-fasting blood glucose ≥11.1 mmol/L, or use of blood glucose-lowering medication. Hypertension was defined as either a measure of systolic blood pressure (BP) ≥ 140 mmHg, diastolic BP ≥90 mmHg or the use of antihypertensive medication. A case of cancer was defined as any type of cancer excluding non-melanoma skin cancer.

**DNA methylation measurements and quality control**

Illumina Infinium Methylation Assay: At the Genetic Laboratory (Department of Internal Medicine, Erasmus University Medical Center, Rotterdam, the Netherlands), genome-wide DNA-methylation levels in 1,613 subjects from the Rotterdam Study were determined using the Illumina HumanMethylation450K BeadChip arrays (Illumina, Inc., San Diego, CA, USA) according to the manufacturers protocol. Genomic DNA was first extracted from whole peripheral blood by standardized salting out methods. Samples were then bisulfite treated using the Zymo EZ-96 DNA-methylation kit (Zymo Research, Irvine, CA, USA), and subsequently hybridized to the arrays.

Preparation and normalization of this array data was performed according to the CPACOR workflow (B. Lehne et al., 2015) using the software package R (www.r-project.org). The idat files were read using the minfi package. Samples with observed technical problems during steps like bisulfite conversion, hybridization, extension or specificity, as well as samples with mismatch between sex of the proband and sex determined by the chr X and Y probe intensities were removed from subsequent analyses. Additionally, only samples with a call rate > 95% per sample were processed further. Probes that had a detection p-value above background (based on sum of methylated and unmethylated intensity values) ≥ 1E-16 in >5% of the samples were removed. This resulted in the remaining of 1,544 samples and 473,682 probes for the analysis. The intensity values as stratified by autosomal and non-autosomal probes, were then quantile normalized for each of the six probe type categories separately: type I methylated red/green, type I unmethylated red/green and type II red/green. Beta values were calculated as proportion of methylated intensity value on the sum of methylated+unmethylated+100 intensities. Blood cell composition of the whole blood samples was estimated using the Houseman method (E. A. Houseman et al., 2012). Beta values being outside of 1.5 times the interquartile range per probe were set to missing. Principal components were calculated from the intensity values of the control probes of all arrays. We used 30 PCs as additional covariates in the association model to adjust for technical bias.

**Epigenome-wide association analysis of mortality:** We conducted time-to-event analyses using Cox proportional hazards models to test for associations between each CpG and mortality outcomes including all-cause death, CVD death and cancer death, using *coxph*() function in the ‘survival’ R library. We considered as confounders socioeconomic, lifestyle and health-related factors. Additional technical covariates were included in the models, such as array number and position on array to account for batch effects during DNA methylation assessment, as well as predicted leukocyte proportions to account for cell heterogeneity.

**Women’s Health Initiative (WHI) – EMPC ancillary study**

**Study participants**: The Women’s Health initiative is a multicenter prospective study of causes of mortality and morbidity among post-menopausal women(Anderson et al., 2003). Between 1993 and 1998, 161,808 women aged 50-79 years were enrolled into the Clinical Trials (n=68,132) or the Observational (n=68,132) arms of WHI. The present study included participants selected into the Epigenetic Mechanisms of PM-Mediated CVD Risk ancillary study (WHI-EMPC), created to elucidate the involvement of epigenetic factors in the relationship between cardiovascular disease susceptibility and airborne particulate matter exposure. The WHI-EMPC is a representative stratified, random sample of 2,200 WHI Clinical Trials participants, and excluded individuals taking anti-arrhythmic medications at the time of the screening visit, or without available buffy coat, core analytes, electrocardiograms, or exposure to ambient air particulate pollution.

**Mortality ascertainment and clinical phenotypes:** Age, height and weight were recorded at study baseline following standard protocols. Questionnaires administered during the WHI screening visit collected participant-reported educational attainment, health behaviors, and medical history including hypertension, prevalent coronary heart disease (cardiac arrest, coronary artery bypass grafting, percutaneous transluminal coronary angioplasty, angina, or myocardial infarction), stroke, heart failure, and cancer. Education was classified as college degree or greater, high school degree or equivalent, or no high school degree or equivalent. Cigarette smoking was dichotomized based on current smoking status. Weekly alcohol intake was estimated in grams per day, based on self-reported alcohol use. Physical activity was quantified in weekly metabolically-equivalent hours estimated from participant self-reports of activities. Body mass index was calculated as kilograms per height in meters squared.

Death occurring until 2010 were ascertained during annual and semi-annual follow-up with participants, family, friends, or medical care providers, and by searches of obituaries and National Death Index records. CVD- and cancer-related deaths were adjudicated based on information provided on death certificates, medical and hospitalization records, and other available records including autopsy reports(Curb et al., 2003).

**DNA methylation measurements and quality control:** Peripheral blood leukocytes were isolated from fasting blood drawn from study participants at study baseline. Genome-wide DNAm at 485,577 CpG sites sites was measured using the Illumina using the Illumina 450K Infinium Methylation BeadChip (Illumina Inc.; San Diego, CA, USA). Methylation was quantified as beta values, indicating the proportion of methylated cytosines at a given CpG site. Methylation data were quality-controlled, excluding CpG sites with detection p-values > 0.01 in over 10% of sample, p-values missing or > 0.01 in over 1% of probes; DNAm values were Beta Mixture Quantile (BMIQ)-normalized to adjust for probe bias(Teschendorff et al., 2013), and adjusted for stage and plate using the empirical Bayes framework implemented in ComBat(Johnson, Li, & Rabinovic, 2007).

**Epigenome-wide association analysis of mortality:** DNAm beta values were rank-normalized, and associations between DNAm at each CpG site and mortality outcomes (all-cause, cvd-related, cancer-related) were modeled using Cox proportional hazards models implemented in the 'survival' R library. Analyses were adjusted for health behaviors, education, body mass index, age, principal components of ancestry calculated from 90 ancestry-informative genetic markers, and prevalent morbidities: coronary heart disease, stroke heart failure, hypertension and cancer,

**Coronary Artery Risk Development in Young Adults (CARDIA) Study**

*Study sample*

The CARDIA study is a prospective multicenter study with 5115 adult, Caucasian and African-American participants ages 18–30 years at recruitment. The recruitment was done from four centers as follows: the total community in Birmingham, AL; selected census tracts in Chicago, IL, and Minneapolis, MN; and from the Kaiser Permanente health plan membership in Oakland, CA. Details of the CARDIA study design have been previously published (Cutter et al., 1991; Friedman et al., 1988). Night examinations were completed beginning with the study initiation in 1985–1986 and in examination years (Y) 0, 2, 5, 7, 10, 15, 20, 25, and 30.

*Adjudication of mortality events*

The CARDIA Study collected data for each participant's death that occurs during the study. Death certificates and/or autopsy reports were requested and reviewed by CARDIA Endpoints Surveillance and Adjudication Subcommittee. Mortality events were obtained from study participants' medical records. For unclear out-of-hospital deaths, the decedent's family, friend, or other person present when the participant died, or the participant's physician was contacted and interviewed to collect sufficient documentation from adjudicating the cause of death. A nurse reviewer/abstractor first reviewed records to rule out obvious non-CVD or death events. Any potential events recorded during this step were then forwarded to an adjudication committee of CARDIA study physicians for verification, as detailed in CARDIA study manual of operations.("CARDIA Endpoint Events Manual of Operations," 2015) All-cause mortality events were ascertained through January, 2020.

*DNA methylation*

Infinium MethylationEPIC BeadChip raw data (IDAT files) were generated from a total of 2,181 blood samples [1,092 at Y20] loaded by the R package *minfi*. Quality control and data preprocessing were conducted using the R package *ENmix* (Xu, Niu, Li, & Taylor, 2016) with default parameter settings. In the quality control step, low-quality methylation measurements were identified by detection p-value <10^-6^ or number of beads <3 (Xu et al., 2016). We excluded 6,209 CpGs with a detection rate <95% and 87 samples with a percentage of low-quality methylation measurements >5% or extremely low intensity of bisulfite conversion probes (less than 3 × standard deviation of the intensity across samples below the mean intensity) (Xu et al., 2016). After excluding low-quality CpGs and samples, we further removed 95 samples that were extreme outliers, as defined by Tukey's method [i.e., <25^th^ percentile – 3 * interquartile range (IQR) or >75^th^ percentile + 3 * IQR] (Tukey, 1977) and based on the average total intensity value [intensity of the unmethylated signal (U) + intensity of the methylated signal (M)] or β value [M / (U + M + 100)] across CpG probes. The remaining samples were preprocessed using ENmix, a model-based background correction method which models methylation signal intensities with a flexible exponential-normal mixture distribution, together with a truncated normal distribution to model background noise (Xu et al., 2016). Dye bias was corrected using *RELIC* (regression on logarithm of internal control probes), which utilizes the intensity values of paired internal control probes that monitor the two-color channels (Xu, Langie, De Boever, Taylor, & Niu, 2017). We then separately quantile-normalized M or U intensities for Infinium I or II probes, respectively. Lastly, low-quality methylation values (detection p-value <10^-6^ or number of beads <3) and extreme β-value outliers across samples (defined by Tukey's method) were set as missing. The final clean methylation working dataset contains 860,627 CpG probes and 957 samples.

**References**

Anderson, G. L., Manson, J., Wallace, R., Lund, B., Hall, D., Davis, S., . . . Prentice, R. L. (2003). Implementation of the Women's Health Initiative study design. *Annals of epidemiology, 13*(9), S5-S17.

Aryee, M. J., Jaffe, A. E., Corrada-Bravo, H., Ladd-Acosta, C., Feinberg, A. P., Hansen, K. D., & Irizarry, R. A. (2014). Minfi: a flexible and comprehensive Bioconductor package for the analysis of Infinium DNA methylation microarrays. *Bioinformatics, 30*(10), 1363-1369. doi:10.1093/bioinformatics/btu049

Aryee, M. J., Jaffe, A. E., Corrada-Bravo, H., Ladd-Acosta, C., Feinberg, A. P., Hansen, K. D., & Irizarry, R. A. (2014). Minfi: a flexible and comprehensive Bioconductor package for the analysis of Infinium DNA methylation microarrays. *Bioinformatics, 30*(10), 1363-1369.

Bell, B., Rose, C. L., & Damon, A. (1966). The Veterans Administration longitudinal study of healthy aging. *The Gerontologist, 6*(4), 179-184.

Bibikova, M., Barnes, B., Tsan, C., Ho, V., Klotzle, B., Le, J. M., . . . Shen, R. (2011). High density DNA methylation array with single CpG site resolution. *Genomics, 98*(4), 288-295. doi:10.1016/j.ygeno.2011.07.007

CARDIA Endpoint Events Manual of Operations. (2015). Retrieved from <http://www.cardia.dopm.uab.edu/images/more/recent/CARDIA_Endpoint_Events_MOO_v03_20_2015.pdf>.

Caspersen, C. J., Bloemberg, B. P., Saris, W. H., Merritt, R. K., & Kromhout, D. (1991). The prevalence of selected physical activities and their relation with coronary heart disease risk factors in elderly men: the Zutphen Study, 1985. *Am J Epidemiol, 133*(11), 1078-1092. doi:10.1093/oxfordjournals.aje.a115821

Chen, Y.-a., Lemire, M., Choufani, S., Butcher, D. T., Grafodatskaya, D., Zanke, B. W., . . . Weksberg, R. (2013). Discovery of cross-reactive probes and polymorphic CpGs in the Illumina Infinium HumanMethylation450 microarray. *Epigenetics, 8*(2), 203-209.

Curb, J. D., Mctiernan, A., Heckbert, S. R., Kooperberg, C., Stanford, J., Nevitt, M., . . . Criqui, M. (2003). Outcomes ascertainment and adjudication methods in the Women's Health Initiative. *Annals of epidemiology, 13*(9), S122-S128.

Cutter, G. R., Burke, G. L., Dyer, A. R., Friedman, G. D., Hilner, J. E., Hughes, G. H., . . . et al. (1991). Cardiovascular risk factors in young adults. The CARDIA baseline monograph. *Control Clin Trials, 12*(1 Suppl), 1S-77S.

Davis, S., Du, P., Bilke, S., Triche, T., & Bootwalla, M. (2017). methylumi: Handle Illumina Methylation Data. R package Version 2.20. 0. In.

Dawber, T. R., Meadors, G. F., & Moore Jr, F. E. (1951). Epidemiological approaches to heart disease: the Framingham Study. *American Journal of Public Health and the Nations Health, 41*(3), 279-286.

Debrabant, B., Soerensen, M., Christiansen, L., Tan, Q., McGue, M., Christensen, K., & Hjelmborg, J. (2018). DNA methylation age and perceived age in elderly Danish twins. *Mechanisms of ageing and development, 169*, 40-44.

Du, P., Zhang, X., Huang, C.-C., Jafari, N., Kibbe, W. A., Hou, L., & Lin, S. M. (2010). Comparison of Beta-value and M-value methods for quantifying methylation levels by microarray analysis. *BMC Bioinformatics, 11*(1), 587. doi:10.1186/1471-2105-11-587

Feinleib, M., Kannel, W. B., Garrison, R. J., McNamara, P. M., & Castelli, W. P. (1975). The Framingham offspring study. Design and preliminary data. *Preventive medicine, 4*(4), 518-525.

Ferrucci, L., Bandinelli, S., Benvenuti, E., Di Iorio, A., Macchi, C., Harris, T. B., & Guralnik, J. M. (2000). Subsystems contributing to the decline in ability to walk: bridging the gap between epidemiology and geriatric practice in the InCHIANTI study. *J Am Geriatr Soc, 48*(12), 1618-1625.

Fortin, J.-P., Labbe, A., Lemire, M., Zanke, B. W., Hudson, T. J., Fertig, E. J., . . . Hansen, K. D. (2014). Functional normalization of 450k methylation array data improves replication in large cancer studies. *Genome biology, 15*(11), 503.

Fried, L. P., Borhani, N. O., Enright, P., Furberg, C. D., Gardin, J. M., Kronmal, R. A., . . . Newman, A. (1991). The cardiovascular health study: design and rationale. *Annals of epidemiology, 1*(3), 263-276.

Friedman, G. D., Cutter, G. R., Donahue, R. P., Hughes, G. H., Hulley, S. B., Jacobs, D. R., Jr., . . . Savage, P. J. (1988). CARDIA: study design, recruitment, and some characteristics of the examined subjects. *J Clin Epidemiol, 41*(11), 1105-1116.

Holle, R., Happich, M., Löwel, H., & Wichmann, H. E. (2005). KORA--a research platform for population based health research. *Gesundheitswesen, 67 Suppl 1*, S19-25. doi:10.1055/s-2005-858235

Houseman, E. A., Accomando, W. P., Koestler, D. C., Christensen, B. C., Marsit, C. J., Nelson, H. H., . . . Kelsey, K. T. (2012). DNA methylation arrays as surrogate measures of cell mixture distribution. *BMC Bioinformatics, 13*(1), 86. doi:10.1186/1471-2105-13-86

Houseman, E. A., Accomando, W. P., Koestler, D. C., Christensen, B. C., Marsit, C. J., Nelson, H. H., . . . Kelsey, K. T. (2012). DNA methylation arrays as surrogate measures of cell mixture distribution. *BMC Bioinformatics, 13*, 86. doi:10.1186/1471-2105-13-86

Ikram, M. A., Brusselle, G., Ghanbari, M., Goedegebure, A., Ikram, M. K., Kavousi, M., . . . Voortman, T. (2020). Objectives, design and main findings until 2020 from the Rotterdam Study. *Eur J Epidemiol, 35*(5), 483-517. doi:10.1007/s10654-020-00640-5

Investigators, A. (1989a). The atherosclerosis risk in communit (aric) stui) y: design and objectwes. *American journal of epidemiology, 129*(4), 687-702.

Investigators, A. (1989b). The atherosclerosis risk in communities (ARIC) study: design and objectives. *American journal of epidemiology, 129*(4), 687-702.

Johnson, W. E., Li, C., & Rabinovic, A. (2007). Adjusting batch effects in microarray expression data using empirical Bayes methods. *Biostatistics, 8*(1), 118-127.

Leek, J. T., & Storey, J. D. (2007). Capturing heterogeneity in gene expression studies by surrogate variable analysis. *PLoS Genet, 3*(9), e161.

Lehne, B., Drong, A. W., Loh, M., Zhang, W., Scott, W. R., Tan, S.-T., . . . Elliott, P. (2015). A coherent approach for analysis of the Illumina HumanMethylation450 BeadChip improves data quality and performance in epigenome-wide association studies. *Genome biology, 16*(1), 37.

Lehne, B., Drong, A. W., Loh, M., Zhang, W., Scott, W. R., Tan, S.-T., . . . Chambers, J. C. (2015). A coherent approach for analysis of the Illumina HumanMethylation450 BeadChip improves data quality and performance in epigenome-wide association studies. *Genome Biology, 16*(1), 37. doi:10.1186/s13059-015-0600-x

Lehne, B., Drong, A. W., Loh, M., Zhang, W., Scott, W. R., Tan, S. T., . . . Chambers, J. C. (2015). A coherent approach for analysis of the Illumina HumanMethylation450 BeadChip improves data quality and performance in epigenome-wide association studies. *Genome Biol, 16*, 37.

Liu, C., Marioni, R., Hedman, Å. K., Pfeiffer, L., Tsai, P., Reynolds, L., . . . Tanaka, T. (2016). A DNA methylation biomarker of alcohol consumption. *Molecular psychiatry*.

Maksimovic, J., Gordon, L., & Oshlack, A. (2012). SWAN: Subset-quantile within array normalization for illumina infinium HumanMethylation450 BeadChips. *Genome biology, 13*(6), R44.

Marioni, R. E., Shah, S., McRae, A. F., Chen, B. H., Colicino, E., Harris, S. E., . . . Cox, S. R. (2015). DNA methylation age of blood predicts all-cause mortality in later life. *Genome biology, 16*(1), 1-12.

Mendelson, M. M., Marioni, R. E., Joehanes, R., Liu, C., Hedman, Å. K., Aslibekyan, S., . . . Yao, C. (2017). Association of body mass index with DNA methylation and gene expression in blood cells and relations to cardiometabolic disease: a mendelian randomization approach. *PLoS medicine, 14*(1), e1002215.

Miller, S., Dykes, D., & Polesky, H. (1988). A simple salting out procedure for extracting DNA from human nucleated cells. *Nucleic acids research, 16*(3), 1215.

Moore, A. Z., Hernandez, D. G., Tanaka, T., Pilling, L. C., Nalls, M. A., Bandinelli, S., . . . Ferrucci, L. (2016). Change in Epigenome-Wide DNA Methylation Over 9 Years and Subsequent Mortality: Results From the InCHIANTI Study. *The Journals of Gerontology: Series A, 71*(8), 1029-1035. doi:10.1093/gerona/glv118

Mortensen, P., Gøtzsche, H., Bøcker Pedersen, C., & Østrup Møller, J. (2006). The Danish Civil Registration System: a cohort of eight million persons. *Dan Med Bull [online], 53*(4), 441-449.

Pedersen, D. A., Larsen, L. A., Nygaard, M., Mengel-From, J., McGue, M., Dalgård, C., . . . Holm, N. V. (2019). The Danish twin registry: an updated overview. *Twin Research and Human Genetics, 22*(6), 499-507.

Pidsley, R., Wong, C. C., Volta, M., Lunnon, K., Mill, J., & Schalkwyk, L. C. (2013). A data-driven approach to preprocessing Illumina 450K methylation array data. *BMC genomics, 14*(1), 293.

Pisani, P., Faggiano, F., Krogh, V., Palli, D., Vineis, P., & Berrino, F. (1997). Relative validity and reproducibility of a food frequency dietary questionnaire for use in the Italian EPIC centres. *International Journal of Epidemiology, 26 Suppl 1*, S152-160. doi:10.1093/ije/26.suppl_1.s152

Price, E. M., Cotton, A. M., Lam, L. L., Farré, P., Emberly, E., Brown, C. J., . . . Kobor, M. S. (2013). Additional annotation enhances potential for biologically-relevant analysis of the Illumina Infinium HumanMethylation450 BeadChip array. *Epigenetics & chromatin, 6*(1), 4.

Rasmussen-Torvik, L. J., Shay, C. M., Abramson, J. G., Friedrich, C. A., Nettleton, J. A., Prizment, A. E., & Folsom, A. R. (2013). Ideal cardiovascular health is inversely associated with incident cancer: the Atherosclerosis Risk In Communities study. *Circulation, 127*(12), 1270-1275.

Raum, E., Rothenbacher, D., Löw, M., Stegmaier, C., Ziegler, H., & Brenner, H. (2007). Changes of cardiovascular risk factors and their implications in subsequent birth cohorts of older adults in Germany: a life course approach. *European Journal of Cardiovascular Prevention & Rehabilitation, 14*(6), 809-814.

Richardson, M. T., Ainsworth, B. E., Wu, H.-C., Jacobs Jr, D. R., & Leon, A. S. (1995). Ability of the Atherosclerosis Risk in Communities (ARIC)/Baecke Questionnaire to assess leisure-time physical activity. *International journal of epidemiology, 24*(4), 685-693.

Schederecker, F., Cecil, A., Prehn, C., Nano, J., Koenig, W., Adamski, J., . . . Thorand, B. (2020). Sex hormone-binding globulin, androgens and mortality: the KORA-F4 cohort study. *9*(4), 326. doi:10.1530/ec-20-0080

Smyth, G. K. (2005). limma: Linear Models for Microarray Data. In R. Gentleman, V. J. Carey, W. Huber, R. A. Irizarry, & S. Dudoit (Eds.), *Bioinformatics and Computational Biology Solutions Using R and Bioconductor* (pp. 397-420). New York, NY: Springer New York.

Soerensen, M., Li, W., Debrabant, B., Nygaard, M., Mengel-From, J., Frost, M., . . . Tan, Q. (2019). Epigenome-wide exploratory study of monozygotic twins suggests differentially methylated regions to associate with hand grip strength. *Biogerontology, 20*(5), 627-647.

Stel, V. S., Smit, J. H., Pluijm, S. M., Visser, M., Deeg, D. J., & Lips, P. (2004). Comparison of the LASA Physical Activity Questionnaire with a 7-day diary and pedometer. *J Clin Epidemiol, 57*(3), 252-258. doi:10.1016/j.jclinepi.2003.07.008

S0895435603003196 [pii]

Svane, A. M., Soerensen, M., Lund, J., Tan, Q., Jylhävä, J., Wang, Y., . . . Deary, I. J. (2018). DNA methylation and all-cause mortality in middle-aged and elderly Danish twins. *Genes, 9*(2), 78.

Taylor, H. L., Jacobs, D. R., Schucker, B., Knudsen, J., Leon, A. S., & Debacker, G. (1978). A questionnaire for the assessment of leisure time physical activities. *Journal of chronic diseases, 31*(12), 741-755.

Teschendorff, A. E., Jones, A., Fiegl, H., Sargent, A., Zhuang, J. J., Kitchener, H. C., & Widschwendter, M. (2012). Epigenetic variability in cells of normal cytology is associated with the risk of future morphological transformation. *Genome medicine, 4*(3), 1-14.

Teschendorff, A. E., Marabita, F., Lechner, M., Bartlett, T., Tegner, J., Gomez-Cabrero, D., & Beck, S. (2012). A beta-mixture quantile normalization method for correcting probe design bias in Illumina Infinium 450 k DNA methylation data. *Bioinformatics, 29*(2), 189-196.

Teschendorff, A. E., Marabita, F., Lechner, M., Bartlett, T., Tegner, J., Gomez-Cabrero, D., & Beck, S. (2013). A beta-mixture quantile normalization method for correcting probe design bias in Illumina Infinium 450 k DNA methylation data. *Bioinformatics, 29*(2), 189-196.

Tobi, E. W., Slieker, R. C., Stein, A. D., Suchiman, H. E. D., Slagboom, P. E., Van Zwet, E. W., . . . Lumey, L. (2015). Early gestation as the critical time-window for changes in the prenatal environment to affect the adult human blood methylome. *International journal of epidemiology, 44*(4), 1211-1223.

Triche Jr, T. J., Weisenberger, D. J., Van Den Berg, D., Laird, P. W., & Siegmund, K. D. (2013). Low-level processing of Illumina Infinium DNA methylation beadarrays. *Nucleic acids research, 41*(7), e90-e90.

Troisi, R. J., Heinold, J. W., Vokonas, P. S., & Weiss, S. T. (1991). Cigarette smoking, dietary intake, and physical activity: effects on body fat distribution—the Normative Aging Study. *The American journal of clinical nutrition, 53*(5), 1104-1111.

Tukey, J. (1977). *Exploratory Data Analysis*: Pearson.

Van Iterson, M., Tobi, E. W., Slieker, R. C., Den Hollander, W., Luijk, R., Slagboom, P. E., & Heijmans, B. T. (2014). MethylAid: visual and interactive quality control of large Illumina 450k datasets. *Bioinformatics, 30*(23), 3435-3437.

White, A. D., Folsom, A. R., Chambless, L. E., Sharret, A. R., Yang, K., Conwill, D., . . . Investigators, A. (1996). Community surveillance of coronary heart disease in the Atherosclerosis Risk in Communities (ARIC) Study: methods and initial two years' experience. *Journal of clinical epidemiology, 49*(2), 223-233.

Xu, Z., Langie, S. A., De Boever, P., Taylor, J. A., & Niu, L. (2017). RELIC: a novel dye-bias correction method for Illumina Methylation BeadChip. *BMC Genomics, 18*(1), 4. doi:10.1186/s12864-016-3426-3

Xu, Z., Niu, L., Li, L., & Taylor, J. A. (2016). ENmix: a novel background correction method for Illumina HumanMethylation450 BeadChip. *Nucleic Acids Res, 44*(3), e20. doi:10.1093/nar/gkv907

Zeilinger, S., Kühnel, B., Klopp, N., Baurecht, H., Kleinschmidt, A., Gieger, C., . . . Illig, T. (2013). Tobacco Smoking Leads to Extensive Genome-Wide Changes in DNA Methylation. *PLOS ONE, 8*(5), e63812. doi:10.1371/journal.pone.0063812

Zhang, Y., Wilson, R., Heiss, J., Breitling, L. P., Saum, K.-U., Schöttker, B., . . . Brenner, H. (2017). DNA methylation signatures in peripheral blood strongly predict all-cause mortality. *Nature Communications, 8*(1), 1-11.
